# Comparing tangible retinal image characteristics with deep learning features reveals their complementarity for gene association and disease prediction

**DOI:** 10.1101/2024.12.23.24319548

**Authors:** Michael J. Beyeler, Olga Trofimova, Dennis Bontempi, José Vargas Quiros, Ilenia Meloni, Bart Liefers, Adham Elwakil, Sacha Bors, Ian Quintas, Leah Böttger, Ilaria Iuliani, Sofia Ortin Vela, Federica M. Conedera, Mattia Tomasoni, Ciara Bergin, Reinier O. Schlingemann, Caroline C.W. Klaver, VascX Consortium, David M. Presby, Sven Bergmann

**Affiliations:** Dept. of Computational Biology, University of Lausanne, Lausanne, Switzerland; Swiss Institute of Bioinformatics, Lausanne, Switzerland; Dept. of Ophthalmology, Erasmus University Medical Center, Rotterdam, The Netherlands; Dept. of Epidemiology, Erasmus University Medical Center, Rotterdam, The Netherlands; Dept. of Ophthalmology, University of Lausanne, Fondation Asile des Aveugles, Jules Gonin Eye Hospital, Lausanne, Switzerland; Platform for Research in Ocular Imaging, Fondation Asile des Aveugles, Jules Gonin Eye Hospital, Lausanne, Switzerland; Department of BioMedical Research, Immunology RIA, Inselspital, University of Bern, Bern, Switzerland; University Medical Centres, Amsterdam, The Netherlands; Dept. of Ophthalmology, Radboud University Medical Center, Nijmegen, the Netherlands; Institute of Molecular and Clinical Ophthalmology, University of Basel, Switzerland; Dept. of Integrative Biomedical Sciences, University of Cape Town, Cape Town, South Africa

## Abstract

Advances in AI, including deep learning (DL), are transforming medical image analysis by enabling automated disease risk predictions. However, DL’s outputs and latent space representations often lack interpretability, impeding clinical trust and biological insight. In this study, we evaluated *RETFound*, a foundation model for retinal images, by comparing its predictive performance and genetic associations to those obtained using clinically interpretable tangible image features (TIFs). Our findings revealed that fine-tuning *RETFound* to predict TIFs provides reasonable estimates for simpler TIFs, like vessel densities (*R*^2^ = 0.91-0.93), but much less accurate approximations for more complex TIFs, like vessel tortuosities (*R*^2^ = 0.25-0.43), highlighting *RETFound’s* limitations to fully characterise the retinal vasculature. We also utilized genome wide association studies on *RETFound*’s latent space, the predicted TIFs, and their measured counterparts to better understand the physiological features that *RETFound* may be focusing on. We find that its latent space variables have many genetic associations, in particular with pathways involved in pigmentation, but only a small overlap with the significant genes identified from measured or predicted TIFs. Analysing the predictive value of the latent space variables, predicted and measured TIFs for clinical endpoints, we find that hybrid models that include all these features perform best for predicting blood pressure and body mass index, indicating that augmenting deep learning models with manually curated features may improve overall prediction capacity. Overall, this study highlights the synergistic potential of integrating deep learning with classical feature extraction, advancing our understanding of retinal biology and disease mechanisms, and paving the way toward improved diagnostic and prognostic tools in ophthalmology.

## Introduction

Medical imaging has become essential for diagnosing diseases and providing presymptomatic risk prognoses. The use of computer-aided analysis, particularly through deep learning (DL), has made these processes not only more accurate but also scalable to large datasets. Recently, foundation models have emerged as a powerful DL approach generating initial models pre-trained on extensive image datasets using self-supervised learning approaches, such as masked autoencoders [1]. Such models can then be applied across a wide range of use cases as they provide a robust and efficient latent space that captures image variability in a low-dimensional form. The basic idea is to use the “encoder” of the model that transforms an image into latent variables (LVs), which can then be used as features for disease identification or prediction. In some cases, the training procedure also involves “fine-tuning” the weights of the encoder, effectively co-adapting the latent space to provide the most pertinent features for the end-point prediction.

While there are ample examples showing the power of using foundation models [1–3], they also have several disadvantages: First, the LVs are typically difficult to interpret. Although there exist techniques to elucidate which regions of an image most strongly contribute to variation in a given LV [4–6], it is generally not obvious what exact properties of the image in these regions are captured. Second, the number of LVs is usually relatively large, typically between 2^7^ and 2^10^, increasing the burden of correcting for multiple hypotheses testing. Finally, there is no guarantee that LVs from masked autoencoders, which are optimised to capture the information needed to reconstruct medical images, efficiently capture disease-relevant signals.

In contrast to DL approaches, clinicians and researchers have been examining images for decades to determine tangible image features (TIFs), such as the size or shape of previously identified anatomical objects, and for many of those there is well-established medical evidence showing their relevance for certain diseases. Recently, automated methods have been increasingly utilized for TIF extraction from medical images, facilitating large-scale analysis and enhancing diagnostic capabilities [7–13]. Among various imaging modalities, colour fundus images (CFIs), obtained via standard color fundus photography of the retina, have emerged as a particularly valuable tool due to their noninvasive nature and ability to predict a wide range of diseases. CFIs provide a cost-effective, in vivo overview of retinal morphology and some information on the underlying choroid and potential pathologies thereof [14], making them instrumental in monitoring ocular diseases such as diabetic retinopathy, macular degeneration, and glaucoma. Furthermore, they enable early risk prediction for systemic conditions, including cardiovascular and cerebrovascular disease, chronic kidney disease, and diabetes [15]. CFIs are also an attractive image modality for investigation because they support the extraction of vascular-based TIFs, facilitated by the availability of public datasets with ground-truth segmentations [16–19]. Of note, we recently published a study of 17 different morphological vascular phenotypes extracted from over 130k fundus images of close to 72k UK Biobank participants. We found that a substantial portion of the variability in these measured TIFs (mTIFs)—features of longstanding interest to ophthalmologists, including median vessel diameter, diameter variability, main temporal angles, vascular density, central retinal equivalents, the number of bifurcations, and tortuosity—can be attributed to genetic factors, with SNP-based heritability estimates ranging from 5% to 25%. Moreover, we observed a large number of genetic association signals with genes and pathways that appear plausible to be involved in modulating the mTIFs, and many significant correlations with both ocular and systemic diseases or their risk factors [13]. However, mTIFs are inherently limited by human-driven feature selection, which dictates what is considered relevant in disease. Additionally, mTIFs may also suffer from poor generalizability in automated settings, as mathematical “one-size-fits-all” heuristics are often employed for feature extraction.

Here we investigate how mTIFs and DL-based features compare against and complement each other in the context of genetics and disease by leveraging *RETFound* [2], a recently developed transformer-based [20] foundation model for retinal images pre-trained using masked image reconstruction. We selected *RETFound* because it is a large-scale foundation model pretrained on extensive retinal imaging data, enabling extraction of generalizable latent features for multiple downstream tasks, including disease prediction and genetic association studies. This distinguishes it from task-specific models trained only for single diseases. To better understand the potential limitations of the information embedded in *RETFound*’s latent space representation, we examine how well it can predict retinal vascular mTIFs. This exploration is driven by the fact that, unlike medical endpoints, mTIFs are entirely determined by the image itself. We utilize genomic analysis to assess the underlying genetic architectures as they can inform upon what physiological aspect an LV represents within an image. To determine if TIFs outperform or complement foundation models in prediction tasks, we build models that include TIFs and LVs and assess their ability to predict disease. Lastly, as classical retinal features are established using heuristic approaches that may fail to generalize, we use *RETFound’s* prediction of mTIFs to derive “deep” TIFs (dTIFs), and compare their genetic architectures as well as disease predictive capacities (see Fig. 1 for a graphical overview of our study). In summary, using *RETFound* and CFIs as an example, our study sheds light on (i) whether foundation models can predict tangible (“classical”) image features, (ii) whether their latent variables can gain interpretability using genomic analyses, and (iii) whether mTIFs and deep features complement in regards to their genetic signals and disease prediction.

**Figure 1.**
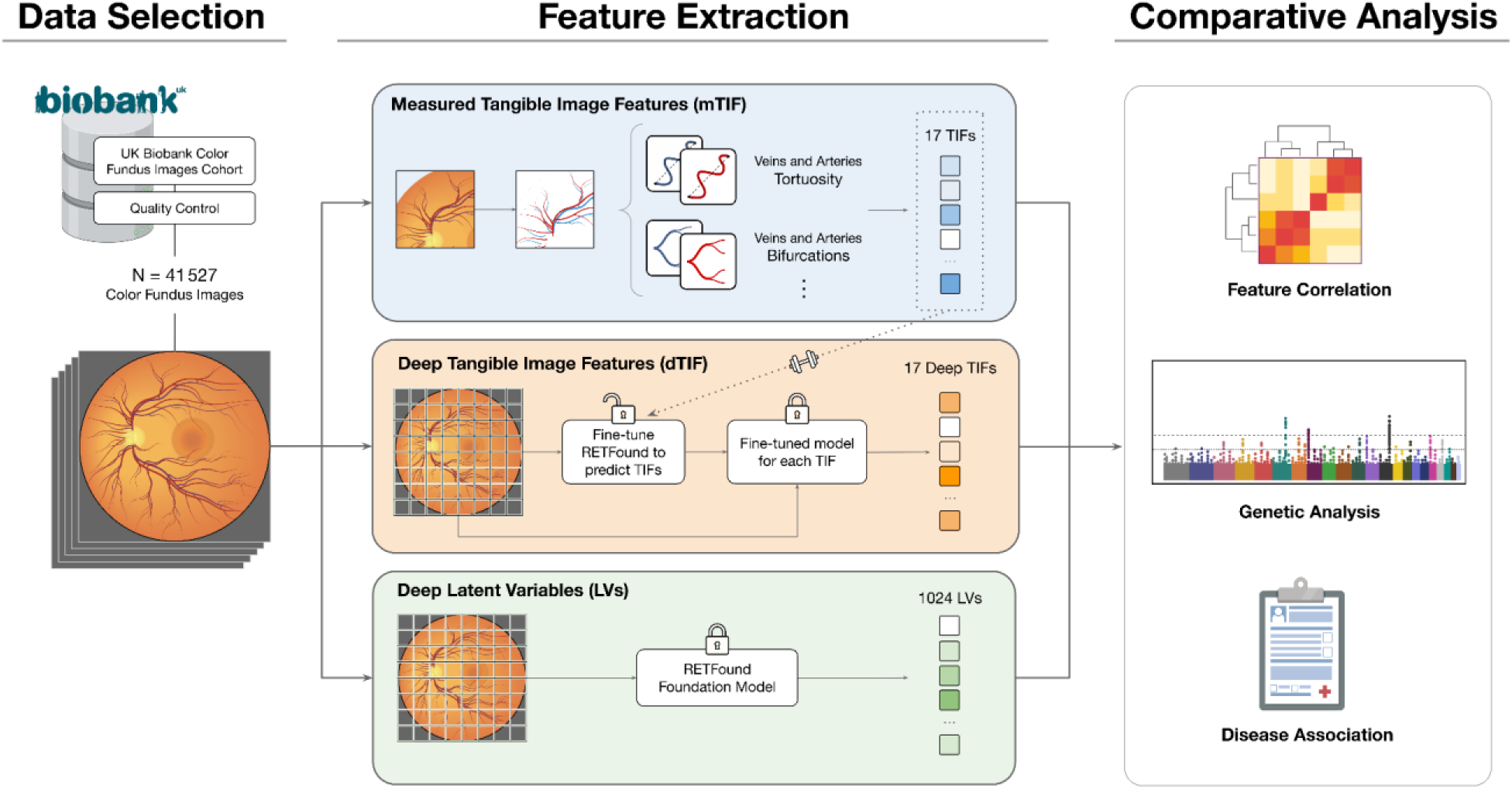
Graphical overview of the study. (a) We selected a subset of genotyped participants from the UK Biobank who had colour fundus images (CFIs) taken, as well as available health outcome information and risk factors. (b) Top panel: Using a previously developed analysis pipeline to characterise the retinal vasculature [13], we extracted tangible image features (TIFs), such as vessel tortuosity or the number of bifurcations, from each CFI. We also used the retinal foundation model *RETFound* [2] as a DL approach to characterise CFIs both (middle) in terms of “deep TIFs” by fine-tuning *RETFound* to predict the 17 measured TIFs and (bottom) by extracting a set of 1 024 latent variables (LVs) using *RETFound’s* pre-trained weights. (c) For downstream comparative analyses, we employed feature correlations, genetic analyses, and disease associations.

## Results

### Phenotypic data generation in terms of vascular TIFs and LVs

Using the methodology described in our previous work [12], we measured 17 TIFs from CFIs of 41 527 genotyped participants from the UK Biobank with complete measurements (see Suppl. Fig. 1 for inclusion-exclusion criteria). Each CFI was also characterised in terms of 1 024 LVs extracted with *RETFound* prior to any fine-tuning, using the weights of the network provided by the authors [2] (see Methods for details).

### Modelling vascular TIFs with *RETFound*

By design, *RETFound’*s LVs capture much of the information contained in CFIs, including the retinal vasculature. Thus, in principle, these LVs should also be informative of our vascular TIFs. In order to evaluate to what extent any individual TIF can be represented by a single LV, we computed all pairwise correlations between LVs and TIFs (shown as a bi-clustered cross-correlation matrix in Fig. 2a). For each TIF, we then picked the LV with the largest squared correlation, which corresponds to the explained variance of a Simple Linear Regression (SLR) model (𝑅^2^_*SLR*_; top bars in Fig. 2b). Our analysis indicates that *individual* LVs can represent a sizable portion of the variability of relatively simple vascular measures, like the vascular densities (𝑚𝑎𝑥 𝑅^2^_*SLR*_ = 0.36). However, this is not the case for vascular TIFs whose measurement is more intricate, where we observe poor correlations between an individual LV and the arterial central retinal equivalent (𝑚𝑎𝑥 𝑅^2^_*SLR*_ = 0.09), tortuosity (𝑚𝑎𝑥 𝑅^2^_*SLR*_ = 0.01), and tortuosity artery/vein (A/V) ratios (𝑚𝑎𝑥 𝑅^2^_*SLR*_ < 0.01). To assess the relationships between LVs, we conducted correlation and principal component (PC) analyses. Only 0.003% of pairwise associations exhibited an R² ≥ 0.7, and 177 PCs were required to explain 95% of the cumulative variance (Suppl. Fig. 2).

**Figure 2.**
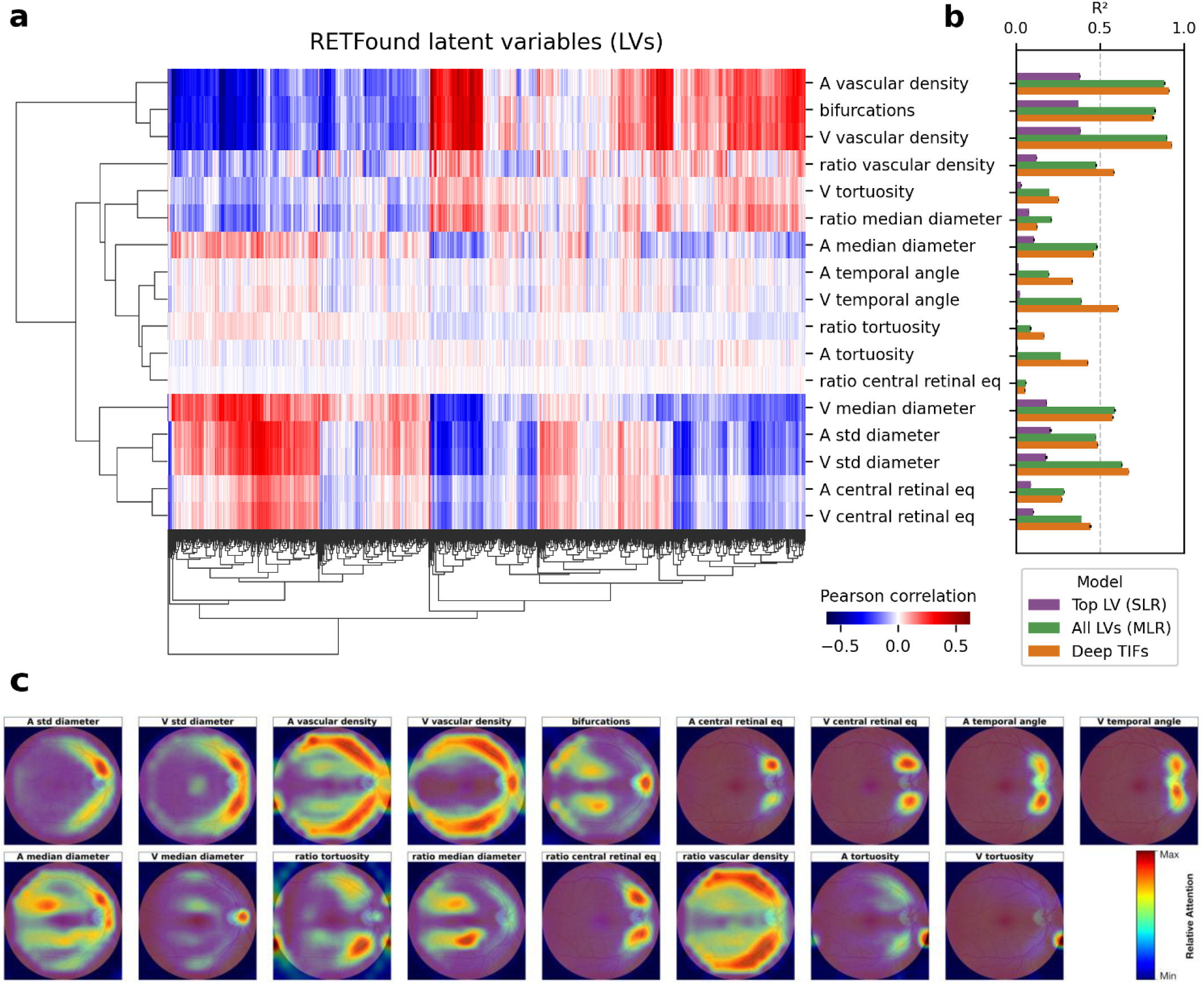
Correlations analysis. (a) Bi-clustered cross-correlation matrix between LVs (x-axis) and TIFs (y-axis). (b) Variance of TIFs explained by the best simple linear regression (SLR) models using the single best baseline LV (purple), multiple linear regression (MLR) models using all baseline LVs (green) and fine-tuning *RETFound* models (orange). Mean out-of-sample R^2^ and their standard errors across the five folds are displayed. A: arterial, V: venous, eq: equivalent, std: standard deviation. (c) Gradient-weighted attention rollout maps for each dTIF, averaged across the cohort, and overlaid on a reference color fundus image. Minimum and maximum attention are calculated independently for each image.

Next, we investigated to what extent combining multiple LVs to predict TIFs can increase the explained variance. As a first step, we trained Multiple Linear Regression (MLR) models with all LVs as features using five-fold cross-validation. The corresponding out-of-sample estimates for the explained variance 𝑅^2^_*MLR*_ for each TIF (middle bars in Fig. 2b) are consistently higher than those from the best SLR model. We observed similar patterns as for SLR, with simple TIFs (such as vascular densities) having more than 90% of their variance explained and much less for more intricate TIFs, requiring explicit identification of vessels and their type. Notably, LVs failed to predict A/V ratios for tortuosity (𝑅^2^_*MLR*_ = 0.09) and central retinal equivalents (𝑅^2^_*MLR*_ = 0.06). In a second step we directly trained *RETFound* to predict TIFs, resulting in 17 fine-tuned *RETFound* models providing TIF estimates as their output, which we call “deep TIFs” (dTIFs). For most of these dTIFs, the out-of-sample estimates 𝑅^2^_*deep*_ for the explained variances (bottom bars in Fig. 2b) are larger than those of the corresponding MLR models. Specifically, the fine-tuned *RETFound* models outperformed the MLR models for 11 out of 17 TIFs, with a mean improvement in 𝑅^2^ of 0.05 (95% CI = [0.01, 0.08], t(16) = 2.45, p = 0.03). We observed the most substantial increases for the temporal angles (arterial: 𝑅^2^_*deep*_ = 0.34, 𝛥𝑅^2^ = 0.14; venous: 𝑅^2^_*deep*_ = 0.61, 𝛥𝑅^2^ = 0.22) and arterial tortuosity (𝑅^2^_*deep*_ = 0.43, 𝛥𝑅^2^ = 0.16), while the improvements for the other TIFs are marginal or even absent. Gradient-weighted attention rollout maps qualitatively indicated that the model attended to expected regions for all dTIFs (i.e., the parts of the vasculature beds that were used to heuristically determine the corresponding mTIF via the automated pipeline; Fig. 2c).

### Comparing the genetic association signals of TIFs and LVs

We previously showed that many of our TIFs have a sizable genetic component, with heritability estimates ranging from 5% for median vessel diameter to >25% for arterial tortuosity [12]. Our genome-wide association study (GWAS) further revealed many plausible genes and pathways for the vast majority of TIFs. These genetic association signals indicate that our TIFs are likely to capture some processes related to vascular maintenance that are partially modulated by genetic variants. As we do not understand what information is captured by LVs—other than that they can only partially represent TIFs— we sought to elucidate their genetic architectures, as these genetic signals can potentially shed light on the image-based features represented by the LVs. To this end, we performed GWAS, heritability estimation, gene and pathway analyses for all 1 024 LVs of the *RETFound* model (before any fine-tuning) and our 17 dTIFs.

At the SNP-level (Fig. 3a–c), the number of significantly associated variants was higher for mTIFs (91 SNPs at a Bonferroni-adjusted threshold of 5/17×10⁻⁸) and dTIFs (88 SNPs at 5/17×10⁻⁸) than for LVs (63 SNPs at 5/1 024×10⁻⁸). Since LVs are not independent we also performed a GWAS on their 17 leading PCs, explaining 66% of their variance, which resulted in n = 28 SNPs with p < 5/17·10^-8^ (Suppl. Fig. 3). Notably, a subset of LVs showed the strongest associations, with an effect size of β = 0.45 and a nominal significance of p < 10⁻³⁰⁰. rs12913832 at the *HERC2* locus was a key driver of this signal. In contrast, 38.8% of loci were shared between mTIFs and dTIFs, with the numbers of exclusive hits being 39 for mTIFs, 37 for dTIFs, and 53 for LVs. Gene-level aggregation using the *PascalX* tool [21,22] confirmed that the genetic architecture of dTIFs more closely resembles that of mTIFs than LVs (Fig. 3d), and while the strongest hits come from LVs (Fig. 3e), dTIFs had more independent gene discoveries (Fig. 3f). Consistently, the overlap of significantly associated genes was 50.4% between mTIFs and dTIFs, but only 12.4% between mTIFs and LVs (Fig. 3f). The most significant gene-level association was observed for the *HERC2*/*OCA2 locus* (p = 3.78×10^-293^ in LV 813), which also influences some TIFs, albeit less significantly (p = 7.4×10⁻²¹ in venous vascular density). Finally, heritability estimates indicated that most LVs have a modest genetic contribution (h² < 0.1 for 77.5% of LVs), with only one LV exhibiting a heritability h² > 0.21 (Fig. 3g). A similar pattern emerged at the gene level, with LVs showing a less polygenic architecture compared with mTIFs and dTIFs (Fig. 3h). Moreover, while genes linked to TIFs were generally specific to only one or two of the 17 phenotypes, those associated with LVs tended to influence multiple traits (Fig. 3i). Genome-wide inflation factors and *Linkage Disequilibrium (LD) Score Regression (LDSR)* intercepts were within acceptable ranges for all feature sets (Suppl. Fig. 4)

**Figure 3.**
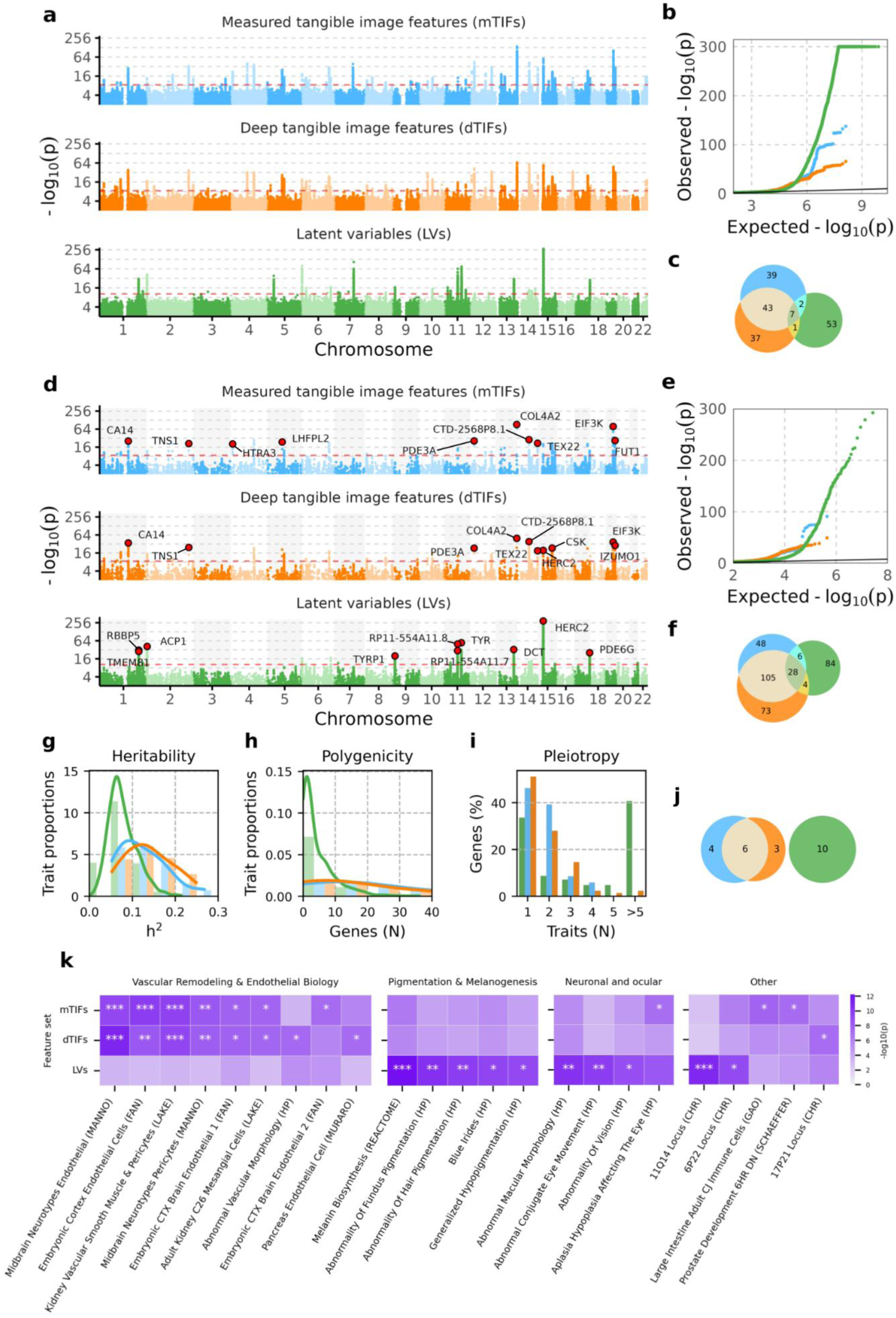
Genetics architectures of mTIFs, dTIFs, and LVs. (a,d) Manhattan plots showing aggregated −log10(p-values) across all traits within each feature set at the SNP level (a) and gene-score level (d). For each genomic position, all p-values observed across traits within a given feature set are shown, enabling visualization of lead loci for the entire feature set in a single panel rather than for a representative trait. Red horizontal lines denote genome-wide Bonferroni significance thresholds adjusted for the number of traits within each feature set. (b,e) Corresponding QQ plots of aggregated SNP-level (b) and gene-level (e) p-values for each feature set. Black lines denote null expectation. (c,f,j) Venn diagrams showing overlap of genome-wide significant SNPs (c) and genes (f), and pathways (j) across feature sets. A SNP, gene, or pathway is considered significant if it reaches genome-wide Bonferroni significance adjusted for the number of traits within each feature set. (g,h) Distributions of heritability (g) and polygenicity (h) across traits within each feature set. The y-axes denote trait proportions (bar values sum to 1 within each feature set), allowing comparison of relative distributions independent of the number of traits. (i) Percentage of genome-wide significant genes (from f) associated with an increasing number of traits within each feature set. (k) Complete list of significant pathways, grouped by functional similarity. Asterisks indicate thresholds of statistical significance (*: α=0.05, **: α=0.01, ***: α=0.001). Genome-wide Bonferroni significance thresholds adjusted for the number of traits within each feature set.

To deepen the interpretability of our genetic results, we performed gene set enrichment testing on the full MSigDB using *PascalX*. The results revealed a complete disjoint of significant pathways between TIFs and LVs (Fig. 3j), and high similarity between mTIFs and dTIFs. An in-depth look at all the significant pathways (Fig. 3k) demonstrated that significant genetic signals from both mTIFs and dTIFs predominantly come from pathways functionally implicated in vascular remodeling and endothelial biology, while genetic signals from LVs mostly point to pigmentation, neuronal, and ocular pathways.

TIF-level analyses (Fig. 4), comparing dTIFs and mTIFs in terms of heritabilities and genetic associations, revealed that heritability estimates for dTIFs were generally comparable to those of the corresponding mTIFs, or in some cases slightly higher (Fig. 4a). Notably, for the arterial temporal angle, the “deep” version showed a substantially higher heritability (h^2^=0.22, 𝛥ℎ^2^ = 0.14). We made a consistent observation at the gene level (Fig. 4b), where dTIFs and mTIFs showed similar numbers of associated genes and large overlaps. Only for venous vascular density, the corresponding arterio-venous ratio, and venous tortuosity the dTIFs had fewer associated genes than their corresponding mTIFs. Interestingly, “deep” arterial tortuosity had 10 additional gene discoveries (10%), despite a slightly lower heritability (𝛥ℎ^2^ = −0.02), whereas its venous counterpart lost 11 discoveries (−32%).

**Figure 4.**
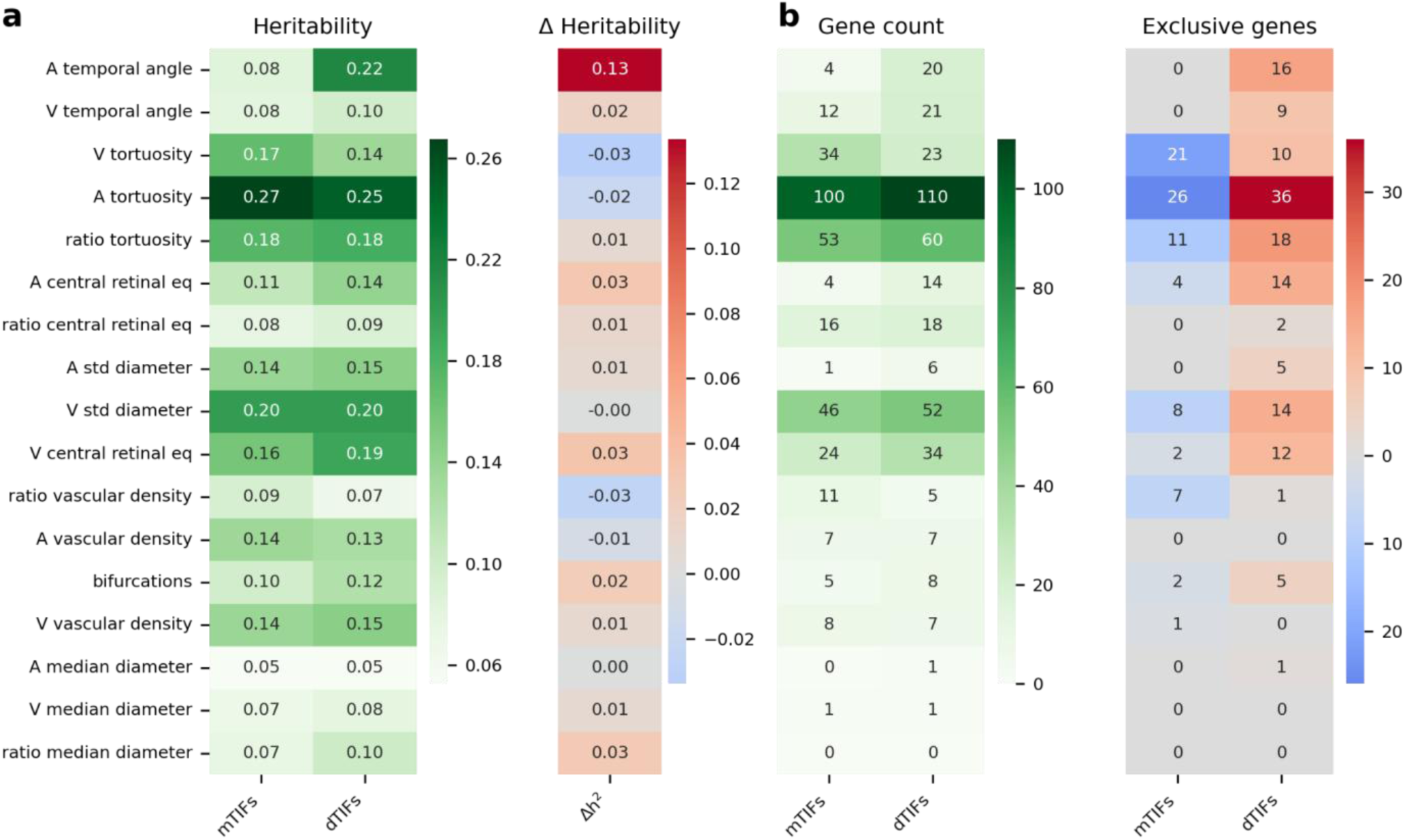
Heritabilities and gene associations per TIF. (a) SNP heritability estimates obtained via *LDSR* and the difference between deep and measured equivalents. (b) Number of associated genes of mTIFs versus dTIFs, including the count of genes exclusive to each (blue: exclusive to mTIFs, red: exclusive to dTIFs). Abbreviations: A = Arterial; V = Venous; std = Standard Deviation; mTIFs = measured Tangible Image Features; dTIFs = Deep Tangible Image Features.

### Associating disease risk with measured and deep TIFs

Next, we investigated to what extent risk factors, ocular and general diseases, and clinical events can be explained by mTIFs or dTIFs. We used linear models that incorporated covariates alongside a single TIF as explanatory variables. We assessed the strength of associations using standardised effect sizes and statistical significance and quantified the total number of unique significant associations across diseases and TIFs to summarise the findings (Fig. 5). Additionally, we applied Fisher’s exact test to assess whether the ratio of significant associations differed between mTIFs and dTIFs. Although not statistically significant, the largest difference in the number of significant diseases tended to be found with the venous temporal angle, with five diseases being significantly associated uniquely with dTIFs (*OR* = 2.74, p = 0.21). While also not significant, diabetes-related eye disease (“diabetes-eye”) tended to display the largest number of differences between TIF types, with five more dTIFs than mTIFs (*OR* = 3.43, p = 0.17). When grouping the diseases into categories, we observed that ocular diseases had significantly more associations only with dTIFs than for mTIFs (*OR* = 1.75, p = 0.02). No such pattern was observed across risk factors (OR = 1.06, p = 0.84) or general diseases and events (OR = 0.92, p = 0.81).

**Figure 5.**
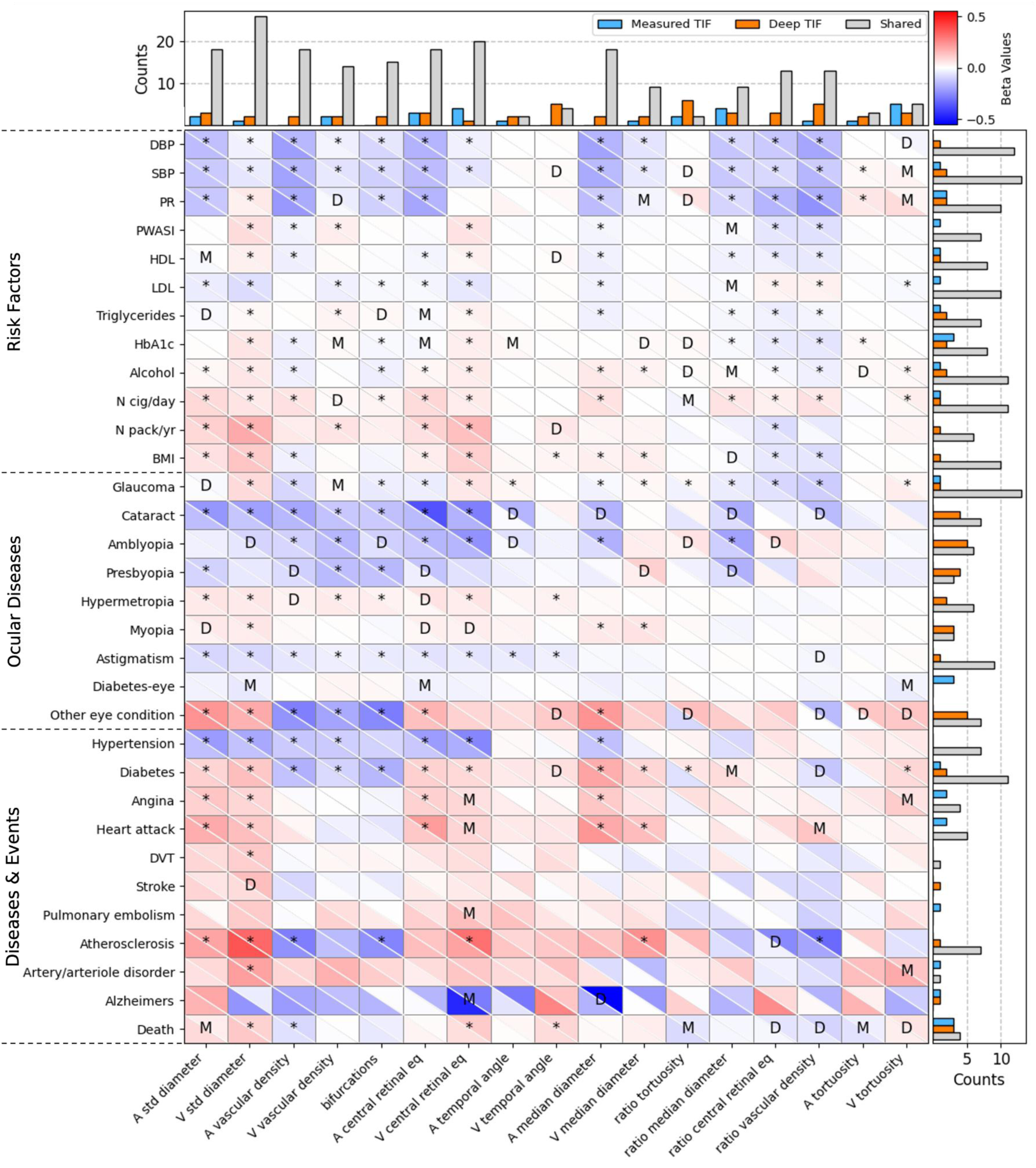
Disease associations with mTIFs and dTIFs. Each cell presents the standardised effects (“beta values”) of a linear model that explains a risk factor, ocular or general disease incident with a single mTIF (lower left portion of a cell) or dTIF (upper right portion of a cell), while accounting for several covariates. Associations significant only for the dTIFs or mTIFs are indicated with ‘D’ or ‘M’, respectively, or a star (*) if significant for both. The bar graphs on the top and right indicate the total number of unique counts for each type of these associations across all diseases and TIFs, respectively. Abbreviations: A = Arterial; V = Venous; DBP = Diastolic Blood Pressure; SBP = Systolic Blood Pressure; Age BP = Age of High Blood Pressure Diagnosis; PR = Pulse Rate; PWASI = Pulse Wave Arterial Stiffness Index; HDL = High-Density Lipoprotein; LDL = Low-Density Lipoprotein; HbA1c = Glycated haemoglobin; N cig/day = self-reported number of cigarettes per day; N pack/yr = self-reported number of packs of cigarettes per year; BMI = Body Mass Index; DVT = Deep Vein Thrombosis.

Finally, we aimed to determine whether models that combine multiple image feature sets increase predictive power over models restricted to just covariates or one feature set at a time. We used regularised multilinear models with eight sets of explanatory features: (i) a baseline model including only covariates (Covar) that were included in each subsequent model; (ii) all 17 mTIFs; (iii) all 17 dTIFs; (iv) the 1 024 RETFound LVs; (v) all mTIFs and dTIFs; (vi) all mTIFs and the 1 024 RETFound LVs; (vii) all dTIFs and the 1 024 RETFound LVs; and (viii) all features. We found that: 1) models that incorporated TIFs with LVs performed best when predicting blood pressure and BMI (Fig. 6a and c); 2) when the outcome was an ocular disease, either there was no additional gain in predictive power over the baseline model, or the LVs tended to outperform the TIFs (Fig. 6b and Suppl. Fig. 5); and 3) both the TIFs and the LVs added to predictive power over the base model for most risk factors and diseases (Fig. 6a-c and Suppl. Fig. 5). Associated statistics for comparing feature predictive capacities can be found in Suppl. Tables 1-2.

**Figure 6.**
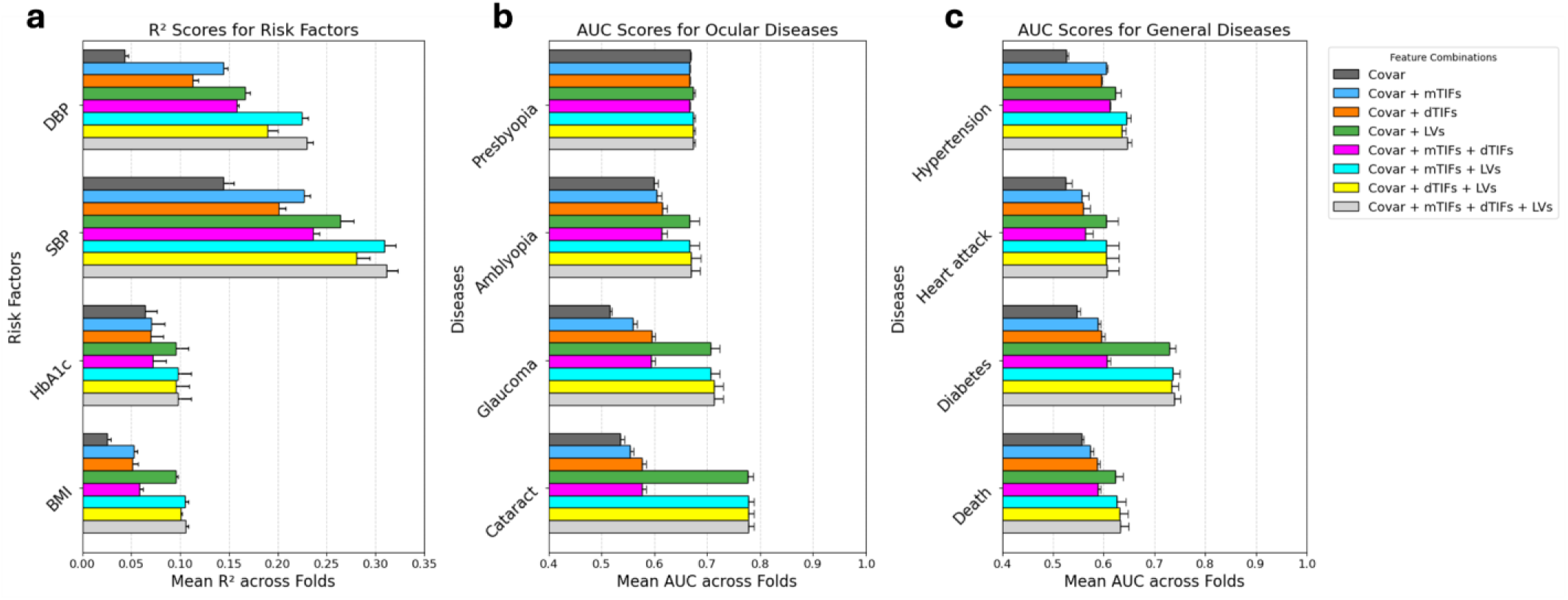
Predictive power of CFI features in assessing risk factors, diseases, and events. (a) Explained variance achieved by linear models built to predict risk factors. (b) Area under the receiver operating characteristic curve (AUC) achieved by logistic regression models built to predict ocular diseases. (c) AUC achieved by logistic regression models built to predict diseases or events. Feature sets include: (i) a baseline model including only covariates (see methods for list), (ii) all 17 mTIFs, (iii) all 17 dTIFs, (iv) 1 024 RETFound LVs, (v) all mTIF and 1 024 RETFound LVs, (vi) all dTIFs and 1 024 RETFound LVs, (vii) all mTIFs and dTIFs, (viii) all features combined. Abbreviations: Covar = Covariates; dTIF = Deep Tangible Image Feature; mTIF = Measured Tangible Image Feature; LVs = Latent Variables; DBP = Diastolic Blood Pressure; SBP = Systolic Blood Pressure; BMI = Body Mass Index; AUC = Area Under the Curve. Error bars reflect 95% confidence intervals.

### Replication in two independent cohorts

We next assessed the reproducibility of our dTIFs, extracting them in two independent cohorts, the Rotterdam Studies (RS, N = 7 205) and the smaller *OphtalmoLaus* (N = 1 714) [23,24]. Specifically, we used the RETFound models we had fine-tuned to predict mTIFs using fundus images from the UKB to generate these “deep TIFs” (dTIFs) from images of the two replication cohorts. We found that the rank order of dTIF predictive capacity across the 17 TIFs was largely preserved in both RS (rho=0.69, p=0.002) and *OphtalmoLaus* (rho=0.66, p=0.004) (Suppl. Fig. 6). Genetically, we found that the heritabilities of dTIFs were highly correlated between the RS and the UK Biobank (r=0.82, p=6e-5; Suppl. Fig. 7). Furthermore, the signed effect sizes of our 88 independent SNP-wise discoveries (Fig. 3c) were overall very consistent in the RS (r=0.87, p=1e-26), and, while less strongly, also in OphtalmoLaus (r=0.70, p=5e-12; Suppl. Fig. 8). Applying the Benjamini-Hochberg procedure at FDR=0.05, we replicated 60 (71%) of the 84 available SNP candidates in the RS. Repeating this procedure at the gene level, we found that replication was even more complete, with 170 (81%) out of 210 gene candidates replicating at FDR=0.05 (Suppl. Fig. 9). We performed individual per-TIF analyses in both the RS and OphtalmoLaus (Suppl. Figs. 10-17), revealing strong candidate replications at both the SNP and the gene level across the majority of TIFs. Lastly, we also observed in the RS that most dTIFs had a larger heritability estimate than their corresponding mTIF (Suppl. Fig. 18; mean difference=0.066 (SE 0.0215), z=3.08, p=0.0021). We report the complete list of replicated SNPs and genes in Suppl. Tables 3-4.

Regarding diseases, we compared the R^2^ of the models designed to predict systolic and diastolic blood pressure using different covariates (Fig. 6a) in the RS replication cohort. The R² values of the models from RS demonstrate a rank order correlation of greater than 0.9 with the corresponding models from the UKBB (RS rho=0.93, p=8.63×10^-4^; OphtalmoLaus rho=0.95, p=2.60×10^-4^), indicating strong reproducibility of these results (Suppl. Fig. 19).

## Discussion

Breakthroughs in computer-aided analysis are revolutionizing the use of medical images for diagnosis and risk assessment. These advances range from supporting physicians in segmenting anatomical structures and extracting precise measurements, to making fully automated disease risk predictions using end-to-end machine learning pipelines. DL algorithms typically hold internal “latent space” representations of the images for this task, but their nonlinear entanglement makes them appear as “black boxes” lacking straightforward explanations. This is problematic in the biomedical domain, where trust by the clinician is essential [25–28]. Furthermore, interpretability is key for deciphering the underlying biological or physiological mechanisms, which is pivotal for advancing treatment [29]. Visual interpretability methods, such as attention maps [4–6] and counterfactual explanations [30], have been developed to address this but often fall short of clarifying exactly how latent variables (LVs) drive predictions (see [28] for some recent advances in tackling this issue).

Here we presented some novel methodological approaches aimed to shed some light on how one can glean insights on what such LVs may present, and in particular what they fall short of. Specifically, our case study investigated *RETFound,* a DL model using the vision transformer architecture and pre-trained using masked image reconstruction that has recently been proposed as a foundation model for retinal images. Our key innovation was to extract *RETFound*’s LVs for a large collection of colour fundus images (CFIs) which we had previously characterised in terms of “tangible image features” (TIFs) of high clinical interpretability [13]. This allowed us to characterise to what extent *RETFound*’s latent space captures those TIFs or could be fine-tuned to predict them. Moreover, the fact that these images stem from genotyped participants with extensive clinical phenotyping enabled us to directly compare the associations of LVs with genotypes and disease states or risk factors with those of TIFs.

Our analysis revealed that, prior to fine-tuning, individual LVs do not strongly correlate with any of our TIFs. Vascular densities and number of bifurcations have the closest LV representation, explaining close to 40% of their variance, while it is less than 20% for the TIFs whose measurement is more intricate. Linear combinations of multiple LVs have substantially higher predictive power for TIFs, in particular for the vascular densities, explaining close to 90% of their variance. Strikingly, fine-tuning *RETFound* to predict TIFs yielded marginal or no improvements compared to the LVs (c.f. [31–33] for similar negative results on fine-tuning in LLMs), with the notable exception of the temporal angles between the main vascular branches and, to a lesser degree, vessel tortuosity. These limited gains from fine-tuning may in part reflect denoising effects or the learning of some relevant proxy trait. Nevertheless, these results suggest that even the advanced transformer DL architecture of *RETFound* and its foundation model parameters trained on more than 900 000 CFIs fail to provide a close approximation of well-established classical ophthalmological parameters of the retinal vasculature. This is remarkable because, in contrast to clinical endpoints, TIFs are fully determined by the images, i.e., they can be extracted through deterministic algorithmic procedures based on conventional image processing.

Our analysis of genetic associations elucidates to what extent *RETFound*’s LVs are modulated genetically and quantifies the overlap with the association signals of the TIFs. We argue that genetic associations support physiological relevance because they point to a causal chain from a DNA difference (i.e. a genetic variant at the level of a single nucleotide) over a molecular variation (such as differential gene expression) to a (typically minute) phenotypic variability. Heritability can be seen as a global summary statistic of such genetic effects, and our results indicate that TIFs predicted by *RETFound*, even when being a relatively poor proxy of their measured counterparts, are nevertheless at least as heritable as the latter, supporting our denoising or relevant proxy hypotheses. Gene-scoring analyses revealed that among *RETFound’*s LVs the strongest association is observed with the *HERC2*/*OCA2* locus, a well-known, strong modulator of pigmentation and eye color [34] that has previously been found as a top hit in a GWAS of LVs from self-supervised deep phenotyping of CFIs [35]. Pathway analyses further supported this observation, with latent variables showing predominant enrichment for pigmentation-related pathways. This indicates that some of the baseline LVs predominantly capture information related to the pigmentation of the retina.

In contrast, dTIFs obtained by fine-tuning *RETFound* demonstrate a genetic architecture that more closely mirrors that of mTIFs. Consistent with this shift, pathway analyses revealed that both mTIFs and dTIFs are enriched for pathways related to vascular remodeling and endothelial biology, in line with their intended representation of retinal vascular morphology. Notably, fine-tuning yielded 73 additional genetic discoveries exclusive to dTIFs, an approximate 50% increase in the number of unique associations observed for mTIFs. Moreover, these genetic signals showed strong replication in an independent cohort, indicating that the fine-tuned model captures stable, transferable features rather than cohort-specific effects. Furthermore, traits such as arterial temporal angles exhibited significantly higher heritability and a greater number of gene discoveries compared to their measured counterparts, suggesting that fine-tuning enhances the capacity of some TIFs to capture subtle yet physiologically pertinent vascular signals. We thus hypothesize that directing *RETFound*’s focus toward TIFs augments to some extent their ability to reflect the underlying vascular biology.

A complementary means of evaluating the pertinence of vascular features is in terms of their capacity to facilitate disease prediction. In this study, we contrasted mTIFs and dTIFs by comparing their standardised effects in logistic and linear models with diseases or risk factors as response variables. We found that the number of significant diseases and risk factors was largely similar between mTIFs and dTIFs, but dTIFs showed more significant associations with ocular diseases compared to mTIFs. This outcome may be attributed to *RETFound’*s initial training on a diverse set of retinal images [2,36]. We hypothesise that its fine-tuned versions, while directed to predict aspects of vascular morphology, might nevertheless retain some of *RETFound’*s broader representation of retinal features. This may allow the model to capture subtle patterns or relationships between vascular morphology and other retinal structures that are relevant to ocular pathologies but not explicitly measured in traditional vascular assessments. The enhanced performance in ocular disease prediction using our fine-tuned models suggests that DL-derived vascular morphology metrics may provide additional informative features for ocular condition detection compared to conventional morphological measurements alone.

Using models that integrate multiple feature types (i.e., baseline, mTIFs, dTIFs, and LVs), we provide evidence in both discovery and replication cohorts that their integration enhances the predictive power for disease. Notably, despite being representative of the entire fundus image, including the vasculature, models that included both LVs from *RETFound* and TIFs (both measured and deep) perform best when predicting blood pressure or BMI, suggesting that TIFs provide complimentary information not fully captured by *RETFound*’s LVs. One potential reason is that the *RETFound* model may not capture the information provided by TIFs is due to its pre-training process through masked autoencoding [37], which is not designed to prescribe particular importance to vascular structures when reconstructing the image. As a result, some critical vasculature information may be excluded from its base weights. Indeed, the original authors of *RETFound* qualitatively report that the model fails to reconstruct much of the finer vasculature present in CFIs. Our results further support this, as we find that fine-tuning *RETFound* to predict vascular TIFs only captures a relatively small portion of their variance, suggesting that the model is unable to learn detailed vascular and vessel type specific information. Similar findings regarding hybrid models that combine manually- and DL-derived features have been observed in other domains, where manually engineered features capture domain-specific knowledge that DL models may overlook when sample sizes are small [38] or when training and test set distributions differ [39,40]. These results emphasize the importance of leveraging both foundation- and expert-derived features to optimize predictive performance. Moreover, future work should examine whether foundation models trained with objectives that explicitly emphasize specific retinal structures better capture and predict these features of interest.

In summary, our findings provide key insights into the interplay between DL-derived features and classical retinal image features in the context of genetics and disease prediction. While *RETFound* can predict certain TIFs, such as vessel densities, with reasonable accuracy, it provides poor estimates of more intricate features, like measures related to artery-to-vein ratios. Despite these limitations, dTIFs exhibit heritability comparable to or greater than their measured counterparts, with a notably large increase in heritability found with temporal angles of the arteries, suggesting that dTIFs may enhance physiologically relevant signal extraction. Furthermore, the genetic and clinical associations of mTIFs and dTIFs show both overlap and complementarity, reinforcing the idea that DL-derived features do not simply replicate classical measurements but may provide additional, distinct insights. Most notably, integrating LVs with TIFs leads to superior predictive performance of cardiovascular risk factors such as SBP and DBP compared to models using either feature set alone, highlighting the potential of hybrid approaches in disease modeling. Overall, this work highlights the synergy between DL and classical feature extraction for advancing our understanding of retinal biology and disease mechanisms, potentially leading to improved diagnostic and prognostic tools in ophthalmology.

## Methods

### Data and quality control

The UK Biobank is a population-based cohort of ∼488k participants with rich, longitudinal phenotypic data, including medical history, and a median 10-year follow-up [41]. Standard retinal 45° CFIs were captured via standard color fundus photography using a Topcon 3D-OCT 1000 Mark II, resulting in a total of 173 814 CFIs from 84 813 individuals. For this study we used 41 527 of these images from the right eye, for which we were able to measure all of our 17 TIFs using our previously described analysis pipeline [13]. 96% of these images passed the quality control threshold we had used in our previous work [13] (see Suppl. Fig. 20). This effectively removed CFIs for which our algorithm failed to identify the optic disk or enough major blood vessels to measure the temporal angles. Since this happens more often for CFIs from older and diseased participants, the cohort of the present study appears to be more “healthy” with respect to its baseline risk factors (see Suppl. Table 5 for a comparison of baseline characteristics to our previous study). Since some of the LVs produced from *RETFound* encoded information specific to eye laterality (left vs. right), we opted to analyze a single eye rather than averaging across both eyes. Genotyping was performed on Axiom arrays for a total of 805 426 markers, from which ∼96 million genotypes were imputed. We used the subset of 15 599 830 SNPs that had been assigned an rsID.

### Measured phenotypes

The 17 mTIFs comprise tortuosity, temporal angles, vessel diameters and diameter variability, vascular density, and the number of bifurcations. With the exception of the number of bifurcations, all of these measurements were performed independently for arteries and veins. To explore asymmetries between arteries and veins, the arterio-venous ratio between diameters, tortuosities and vascular densities were also included. All mTIFs were derived using an automated image analysis pipeline described in our recent publication [13], which follows a three-step procedure: (1) deep learning–based segmentation of retinal vessels from raw color fundus images, (2) reconstruction and classification of vessel centerlines into arteries and veins, and (3) quantitative extraction of morphometric features yielding the final mTIFs.

### Deep learning model

We utilised *RETFound [2]*, a novel foundation model that leverages Google’s vision transformer large patch 16 architecture [42]. Briefly, *RETFound* uses a masked autoencoder (MAE) approach [37] to optimise the model towards reconstructing retinal images. For building *RETFound*, 75% of the image patches were masked and the model was trained to reconstruct the missing patches, thereby learning effective representations of the images provided as training data. We used the encoder portion of this model to extract LVs and make predictions.

### Extracting Latent Variables

To extract latent variables (LVs), we utilized the encoder portion of the pre-trained *RETFound* foundation model. Based on the Vision Transformer Large (ViT-L/16) architecture, the model consists of 24 Transformer blocks with a native embedding dimension of 1 024. Input images were resized to 224×224 pixels and normalized using ImageNet-derived means and standard deviations. During extraction, unmasked 16×16 patches were projected into feature vectors and processed through the encoder’s multi-headed self-attention layers and multilayer perceptrons. The resulting 1 024-dimensional feature vectors were then utilized as LVs for all subsequent downstream genetic and phenotypic analyses.

### Predicting tangible image features

For predicting TIFs, we used the encoder portion of *RETFound* and added a multi-layer perceptron to the head of the model. We utilised the base hyperparameters established in the original *RETFound* manuscript [2] and trained the model over 50 epochs, with a “warm-up” in the 10 first epochs (with the learning rate *r* monotonically increasing from 0 to 5 × 10^−4^), followed by a cosine annealing schedule (where *r* decreases from 5 × 10^−4^ to 1 × 10^−6^). We set the batch size to 16, the dropout rate to 0.2, and the image size to 224×224 pixels. After each epoch, we evaluated the model on a validation dataset and then kept the model that had the lowest mean absolute error across all epochs. For each of the 17 TIFs, we trained separate models 5-fold cross-validation with an effective split of 60% for training, 20% for validation, and 20% for testing in each fold for both RETFound fine-tuning and the multiple linear regression baselines, and report the mean test-set r² across the five folds.

### Covariate corrections

Genetic analysis and disease prediction models were corrected for a set of covariates known to confound phenotypic variability in general, or in the eye specifically. They comprise sex, age, age-squared, sex-by-age, sex-by-age-squared, spherical equivalent (defined as 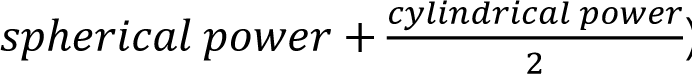), spherical equivalent-squared, imaging instance, assessment centre, genotype measurement batch, and the first 20 genomic PCs (see Suppl. Fig. 21).

### Attention maps

To characterise the model’s focus during inference, we generated attribution maps for all 17 fine-tuned *RETFound* models using a modified gradient-weighted attention rollout [43]. For each input image, we registered hooks on all transformer blocks to retrieve the post-softmax attention matrices and their corresponding gradients with respect to the model output. In each block, and for each attention head, we computed a gradient-weighted attention tensor by taking the element-wise product between the attention weights and their gradients, followed by a ReLU to retain only positive signal. These tensors were then averaged across heads within each block, after which we applied rollout [44]: at every layer we added an identity matrix to account for residual connections, and row-normalized the resulting matrix before multiplying it into the cumulative rollout matrix. This produced a single NxN relevance matrix per image, where N=197 denotes the number of tokens. We focused on the “CLS→patch” attention by extracting the row corresponding to the class token [45], yielding a 14×14 spatial attribution map once reshaped. For each model, we computed attribution maps for all images in the cohort, averaged them to visualise population-level patterns of model focus, and upsampled the resulting matrices to the original image resolution using bicubic interpolation for overlay on a reference colour fundus image.

### Genomic analyses

For GWAS, raw phenotypes were transformed using rank-based inverse normal transformation. GWAS were conducted using *BGENIE [46]*. Only SNPs with minor allele frequency larger than 0.01 and imputation quality larger than 0.8 were kept for downstream analyses. SNP-specific heritabilities and genetic correlations between traits were calculated using *LDSR [47]* which fits an ordinary least squares regression of SNP LD scores against their average chi-squared statistics. *LDSR* considered only common SNPs (MAF > 0.01) with an imputation quality > 0.9. Genome-wide significant SNPs were LD-pruned using LDlinkR::SNPclip (population = GBR; r² < 0.1) to retain an approximately independent set of variants for reporting. Gene and pathway scores were computed using *PascalX* [21,22,48]. Both protein-coding genes and lincRNAs were scored using the approximate “saddle” method, incorporating all SNPs within a 50 kb window surrounding each gene. All pathways from MSigDB v7.2 [49–51] were scored using *PascalX’*s ranking mode, merging and rescoring co-occurring genes separated by less than 100 kb. Among nested subgroups of pathways, we retained only the element with the highest significance. *PascalX* utilises LD structure for accurate computation of gene scores, which in these analyses was derived from the UK10K (hg19) reference panel [52]. For annotating genes in the Manhattan plots, the top ten genes for each feature set (i.e., mTIFs, dTIFs, or LVs) were selected within a 1 000 SNP window.

### Disease associations

Three distinct groups of disease traits were included in this work. First, a set of general systemic risk factors, second, a set of ocular diseases, and third, a set of general diseases and events (see Suppl. Fig. 22). All disease data were collected from the UK Biobank (see Ref. [13] for how we mapped the data field identifiers to each disease included in this study).

To extract beta coefficients, in the case of the continuous risk factors, linear regression was used to estimate standardised effects. For both ocular and general diseases, time-to-event data were binarised, and logistic regression was applied.

When examining the predictive capacity of different feature sets, linear regression was used for continuous risk factors and logistic regression was used for disease prediction. To assess model generalization and obtain robust estimates of predictive performance, five-fold cross-validation was employed, with an 80% training and 20% test split in each fold. Models were trained exclusively on the 80% training portion within each fold, and performance was evaluated on the held-out 20% test set. Reported results correspond to averages across all test folds, such that each subject appears exactly once in a test set, reducing dependence on any single data partition. Both linear and logistic regression models were regularized using elastic net penalties, with optimal values for alpha and the L1 ratio determined by grid search within each fold. We utilized the coefficient of determination (R^2^) as the performance metric for continuous risk factors and area under the receiver operating characteristic curve (AUC) as the performance metric for diseases. To ensure that the predictive capacity of retinal features was not confounded by known risk factors present in the covariates, we created a matched cohort for each binary disease analysis. We downsampled the larger control group by matching each case with a unique control based on the most similar covariate profile, a process performed using optimal bipartite matching (the Hungarian algorithm), ensuring that no control was used more than once. This matching procedure, which resulted in a balanced sample size for cases and controls, allows for a more direct assessment of the independent contribution of the image-derived features. To prevent poor feature-to-sample-size ratios, diseases and events were required to have at least 1 000 samples (cases and controls combined) to be analyzed.

For all regression analyses, traits were corrected for the covariates listed above. To determine the significance of regression coefficients and prediction power, p-values were computed, and the Benjamini-Hochberg procedure was applied using a false discovery rate of 0.05.

### Replication in the Rotterdam Study and OphtalmoLaus

Replication was performed in two cohorts, (1) the Rotterdam Study (RS) [53], and (2) OphtalmoLaus [23], a sub-cohort of CoLaus [54] and a member of the Global RETFound initiative [55]. Replication analyses closely followed our previously described replication pipeline [13] including its Supplementary Methods, with only the following minor changes. In the RS, (1) the number of subjects in the four sub-cohorts available was 5 227 (RS-I), 2 256 (RS-II), 3 166 (RS-III), and 2 229 (RS-IV); (2) the included covariates were age, sex, spherical equivalent, imaging device, image quality (AI score), genomic PCs 1–10, squared terms and interactions (age squared, spherical equivalent squared, age times sex).

For the Rotterdam CFIs, the original dataset comprised 262 965 color fundus images acquired from multiple participants using several different imaging devices. To ensure data quality and consistency, we applied a two-stage filtering pipeline. In the first stage, we retained only macula-centered images, excluded those with missing quality scores, applied an automated image-quality threshold of ≥0.5, and only retained right-eye images for consistency, reducing the dataset from 262 965 to 24 941 images (90.5% reduction). In the second stage, a single highest-quality image was selected per participant using a hierarchical prioritization strategy. Images from the TOPCON 3D OCT-2000 FA Plus device were preferred when available, and among these, the image with the highest quality score was selected. The same criteria were applied across devices when the preferred model was unavailable. The final dataset comprised 8,755 images, with the following device distribution: TOPCON 3D OCT-2000 FA Plus (n=7 168; 81.9%), TOPCON 3D OCT-1000 MARK II (n=1 030; 11.8%), TOPCON 3D OCT-1000 (n=316; 3.6%), TOPCON DRI OCT Triton Plus (n=213; 2.4%), TOPCON TRC-50VT (n=23; 0.3%), and TOPCON DRI OCT Triton (n=5; 0.1%). Regarding OpthalmoLaus, 6 503 images from 2 276 subjects were available, of which 4 890 images from 1 715 subjects passed QC and were used for analyses.

For feature extraction, we used our 17 *RETFound* models that were fine-tuned in the UK Biobank to independently extract dTIFs in each of the two replication cohorts. For genetic replication, candidates at the SNP-level were defined as independent genome-wide-significant loci passing Bonferroni correction adjusted for both the number of independent SNPs and the number of traits (5e-8/17). Candidates at the gene level were defined as PascalX gene scores passing the nominal Bonferroni-corrected threshold adjusted for both the number of tested genes (25819) and the number of traits (17), p_thres=1.14e-7.

## Data availability

GWAS summary statistics, gene and pathway scores, and all data underlying figures will be made available on Zenodo upon publication. GWAS summary statistics will further be made available on the GWAS Catalog. Raw UK Biobank data are protected and not open access; however, they can be obtained upon project creation and acceptance via their platform (https://www.ukbiobank.ac.uk/). mTIFs [13] are already available via this channel; dTIFs and baseline LVs will be added to the platform upon publication.

## Code availability

Code is going to be made available on Zenodo upon publication.

## Data Availability

Genetic and phenotypic data are available upon application to the UK Biobank.
GWAS sumstats and deep-learning models are available upon reasonable request.

https://www.ukbiobank.ac.uk/

