## Supplementary information for "Comparing tangible retinal image characteristics with deep learning features reveals their complementarity for gene association and disease prediction"

### Supplementary Tables

Supplementary Tables

**Supplementary Table 1 | Pairwise comparisons across feature sets for linear regressions used to predict disease.**

**Supplementary Table 2 | Pairwise comparisons across feature sets for logistic regressions used to predict disease.**

**Supplementary Table 3 | List of replicated SNPs. Candidate SNPs replicated jointly, and for each dTIF separately. Reported as rsIDs.**

**Supplementary Table 4 | List of replicated genes. Candidate genes replicated jointly, and for each dTIF separately. Reported as gene symbols.**

**Supplementary Table 5 | Baseline characteristics.** Comparison of covariates and risk factors between our study and complete UK Biobank fundus cohort. Two-sample t-tests were applied to compare continuous variables, tests of proportion were applied to compare binary variables. Note that CFIs from older and diseased subjects were more prone to be excluded, indicating that image quality is not independent from age and disease status.

### Supplementary Figures

#### 1. Inclusion-exclusion criteria

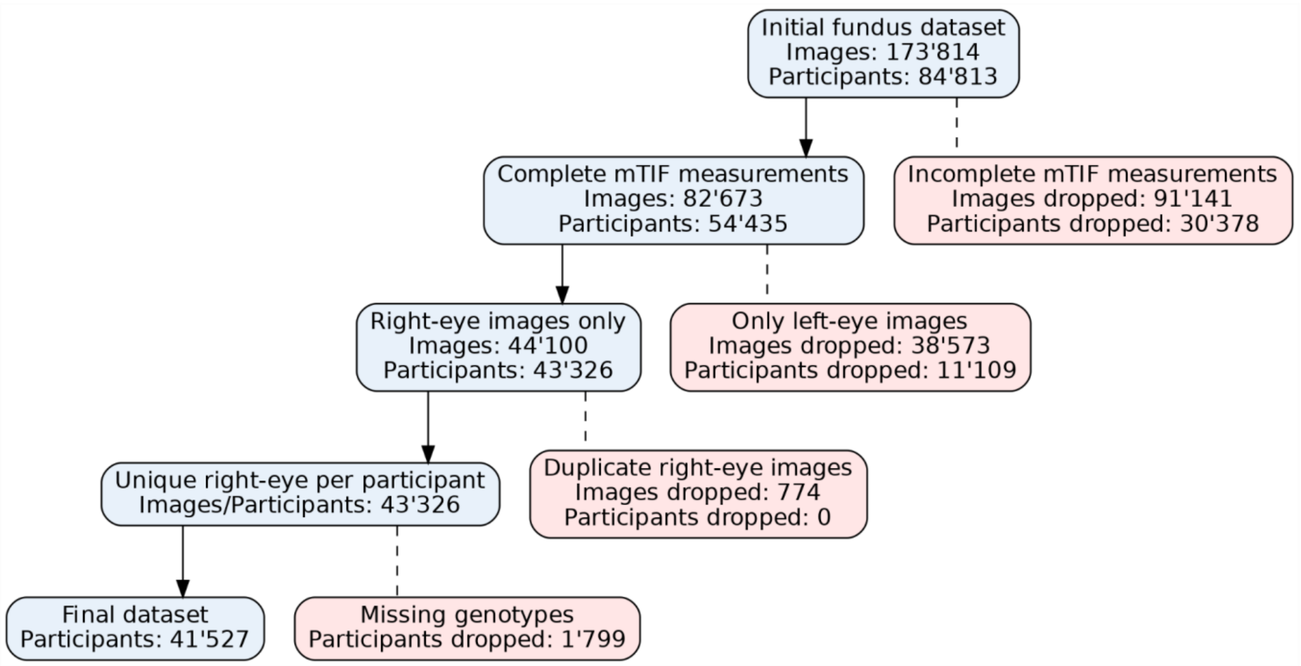

**Supplementary Figure 1 | Inclusion-exclusion flowchart for UK Biobank fundus dataset.** From the initial fundus dataset, images/participants were excluded due to incomplete mTIF measurements and when only left-eye images were available. Analyses were restricted to a single, unique right-eye image per participant and to participants with available genotypes, yielding a final dataset of 41’527 participants. Boxes report image- and participant-level counts at each step.

#### 2. Analysis of *RETFound*'s baseline LVs and their leading PCs

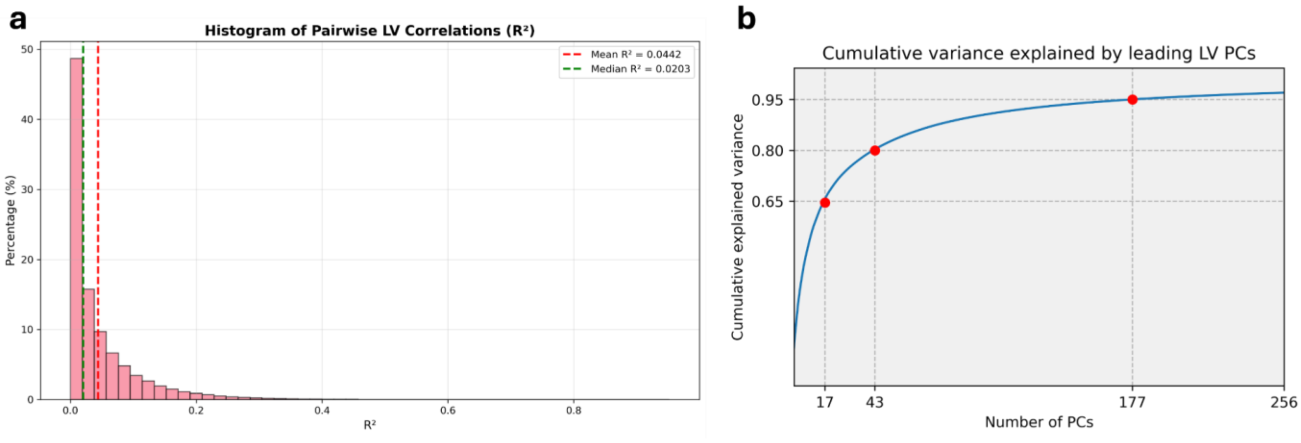

**Supplementary Figure 2 | Associations amongst LVs. a** Histogram displaying the distribution of $R^2$ when correlating all LVs. **b** Cumulative variance explained by leading LV principal components, taken by applying principal component analysis to *RETFound*'s baseline LVs, and showing the cumulative variance explained by the resulting leading PCs.

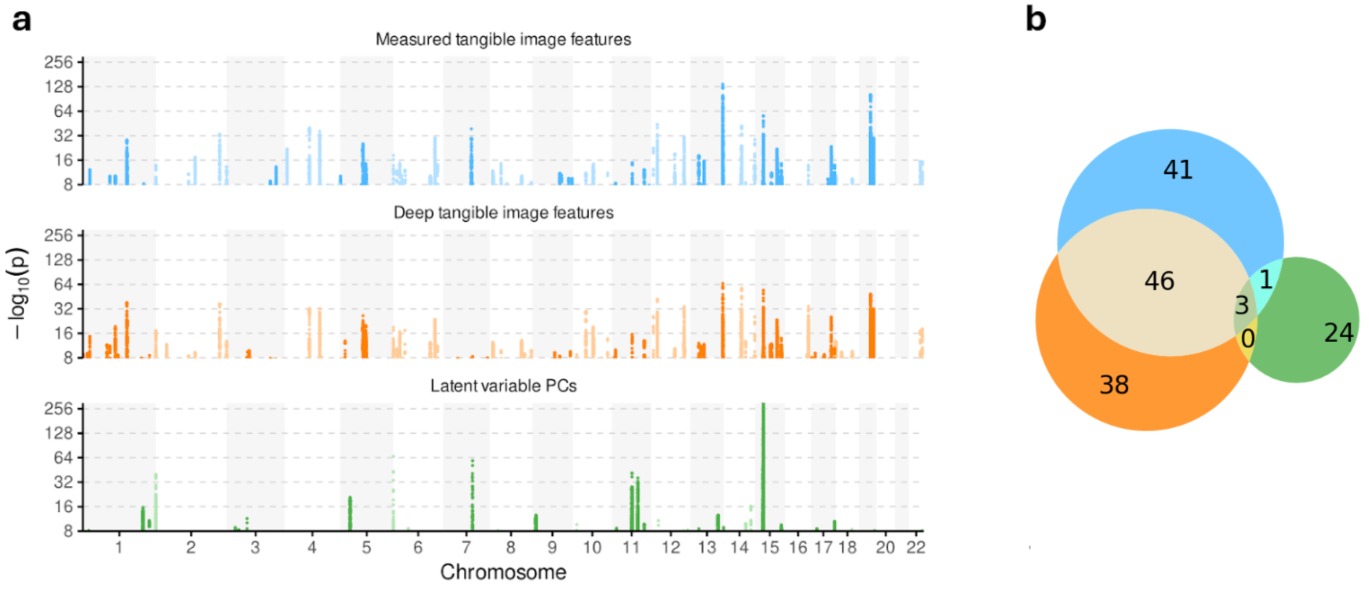

**Supplementary Figure 3 | SNP-wise genetic hits of mTIFs, dTIFs, and leading 17 LV PCs. a** Manhattan plot showing SNPs with nominal Bonferroni-corrected genome-wide significance at $p=5*10^{-8}$. **b** Venn diagram comparing overlap and unique, pruned, SNP-wise discoveries for each method. Significance threshold for each method was set at $p=5*10^{-8} / 17$, 17 being the number of features included in each method. Color codes: Blue: mTIFs, orange: dTIFs, green: LV PCs

#### 3. GWAS quality control

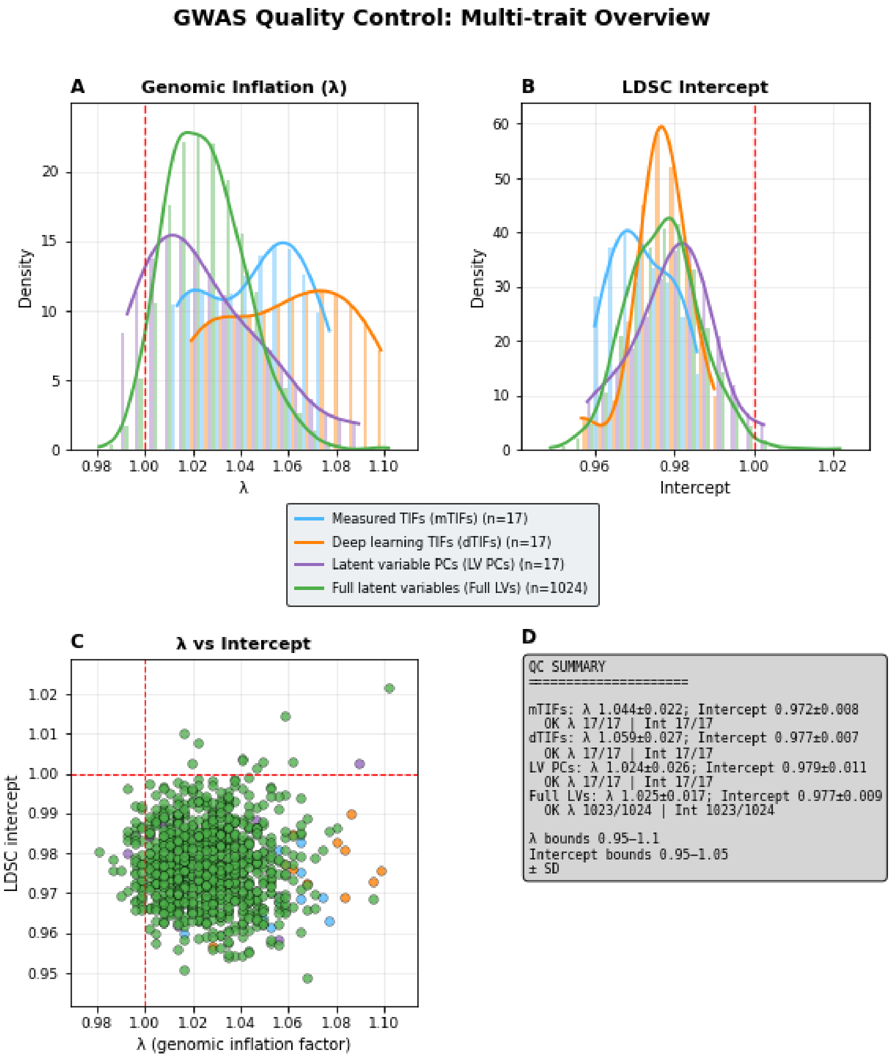

**Supplementary Figure 4 | GWAS quality control metrics for all feature sets.** (A) Distribution of genomic inflation factors (λ) for GWAS for all feature sets. Kernel density estimates are shown for each set. (B) Corresponding distribution of LD Score Regression (LDSC) intercepts. (C) Scatter plot of λ vs LDSC intercept for all GWAS. In (A-C), dashed lines correspond to the expected null values. (D) Statistical summary confirming that all analyzed feature sets satisfy stringent GWAS quality-control criteria, with no evidence of problematic inflation.

#### 4. Predictive capacity of CFI features expanded

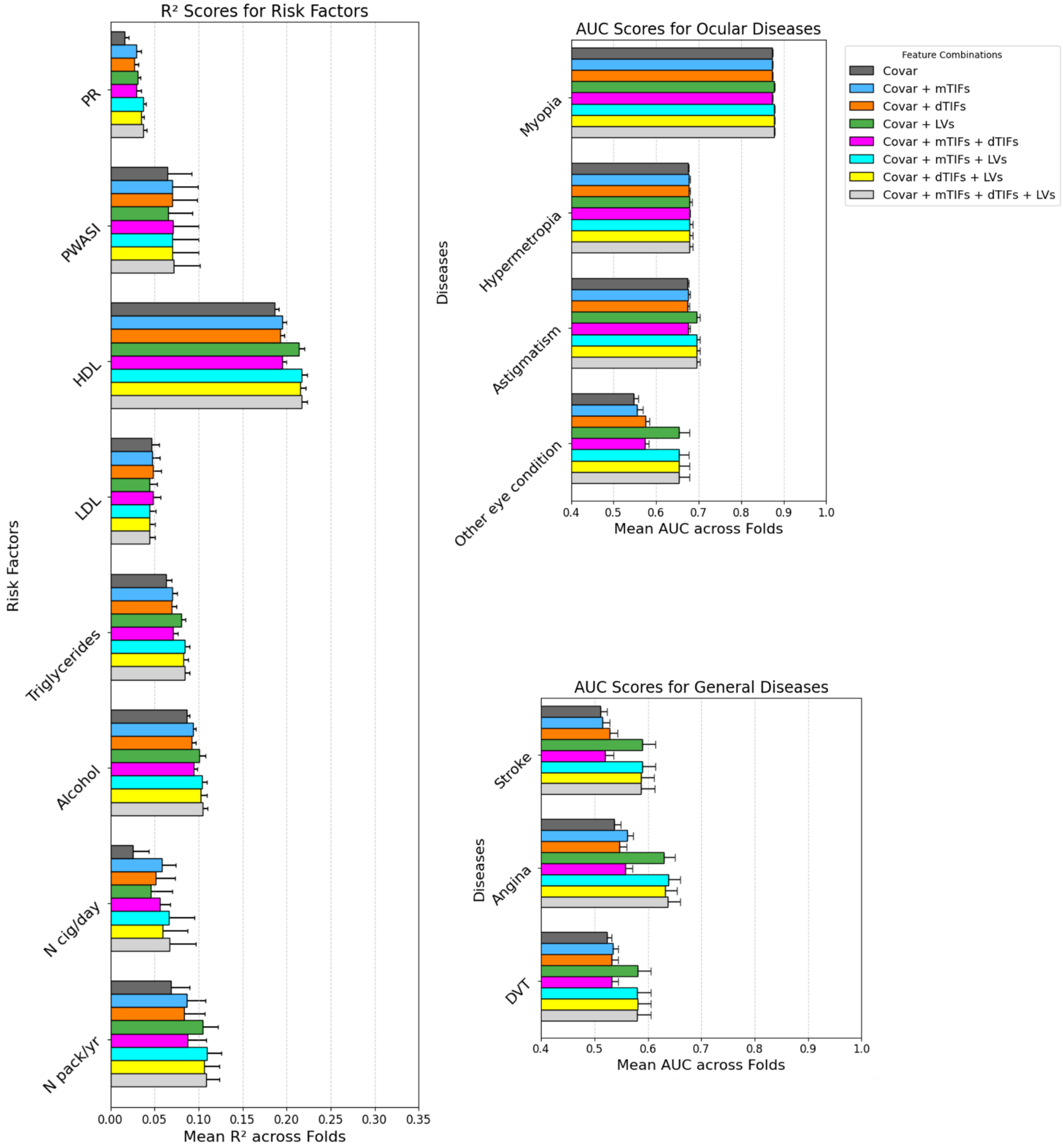

**Supplementary Figure 5 | Predictive capacity of CFI features in assessing risk factors, diseases, and events.** (a) Explained variance achieved by linear models built to predict risk factors. (b) Area under the receiver operating characteristic curve (AUC) achieved by logistic regression models built to predict ocular diseases. (c) AUC achieved by logistic regression models built to predict diseases or events. Feature sets include: (i) a baseline model including only covariates (see methods for list), (ii) all 17 mTIFs, (iii) all 17 dTIFs, (iv) from 1024 RETFound LVs, (v) all mTIF and 1024 RETFound LVs, (vi) all dTIFs and 1024 RETFound LVs, (vii) all mTIFs and dTIFs, (viii) all features combined. Abbreviations: Covar = Covariates; dTIF = Deep Tangible Image Feature; mTIF = Measured Tangible

Image Feature; LV = Latent Variable; AUC = Area Under the Curve; PR = Pulse Rate; PWASI = Pulse Wave Arterial Stiffness Index; HDL = High-Density Lipoprotein; LDL = Low-Density Lipoprotein; N cig/day = self-reported number of cigarettes per day; N pack/yr = self-reported number of packs of cigarettes per year. Error bars reflect 95% confidence intervals.

### 5. Replication in OphtalmoLaus and Rotterdam Study

#### 5.1 Predictions of mTIFs

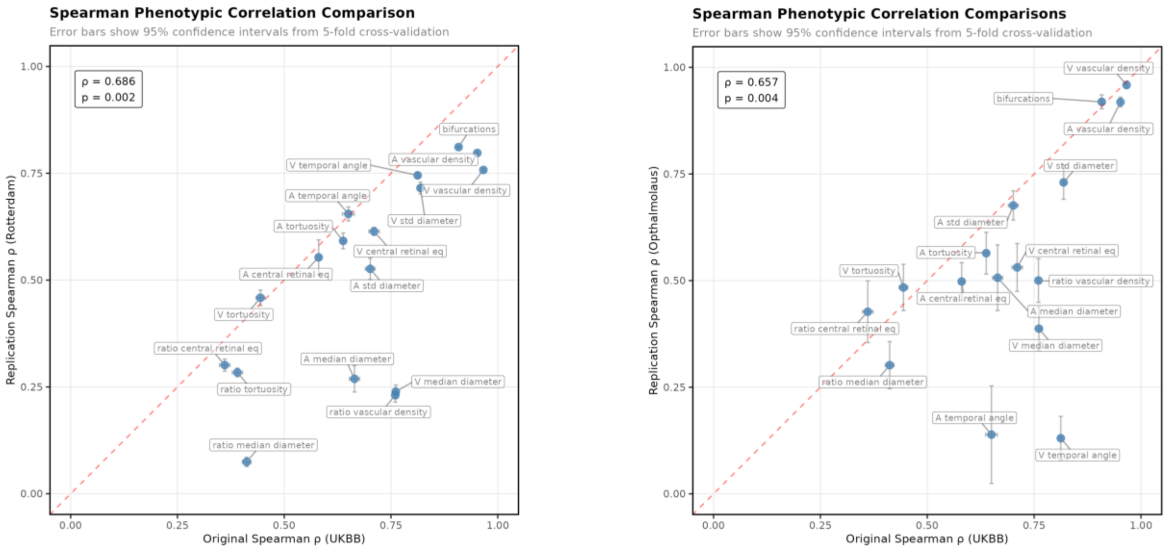

**Supplementary Figure 6 | Predicting mTIF from dTIFs in replication cohorts.** Both panels plot the performance of *RETFound* in predicting mTIFs in the UK Biobank (x-axis) against performance in independent replication cohorts (y-axis; left: Rotterdam Study, right: OphtalmoLaus), quantified by the coefficient of determination ($R^2$). Spearman rank correlations were used to assess concordance in predictive performance ($R^2$) across studies.

#### 5.2 SNP-wise heritabilities of dTIFs

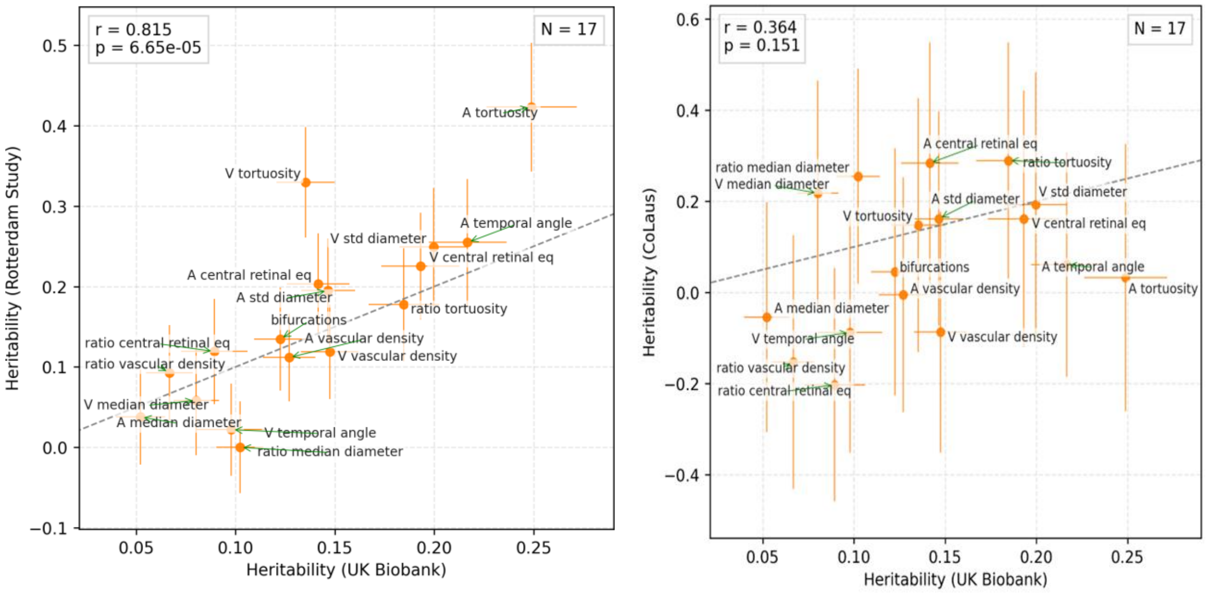

**Supplementary Figure 7 | dTIF heritabilities.** Scatter plots comparing SNP-based heritability estimates of deep tangible image features (dTIFs) in the UK Biobank (x-axis) versus independent replication cohorts (y-axis; left: Rotterdam Study, right: OphtalmoLaus / CoLaus). Error bars denote standard errors of *LDSR* heritability estimates. Spearman correlation coefficients and associated p-values are shown.

#### 5.3 Joint candidate SNP replication

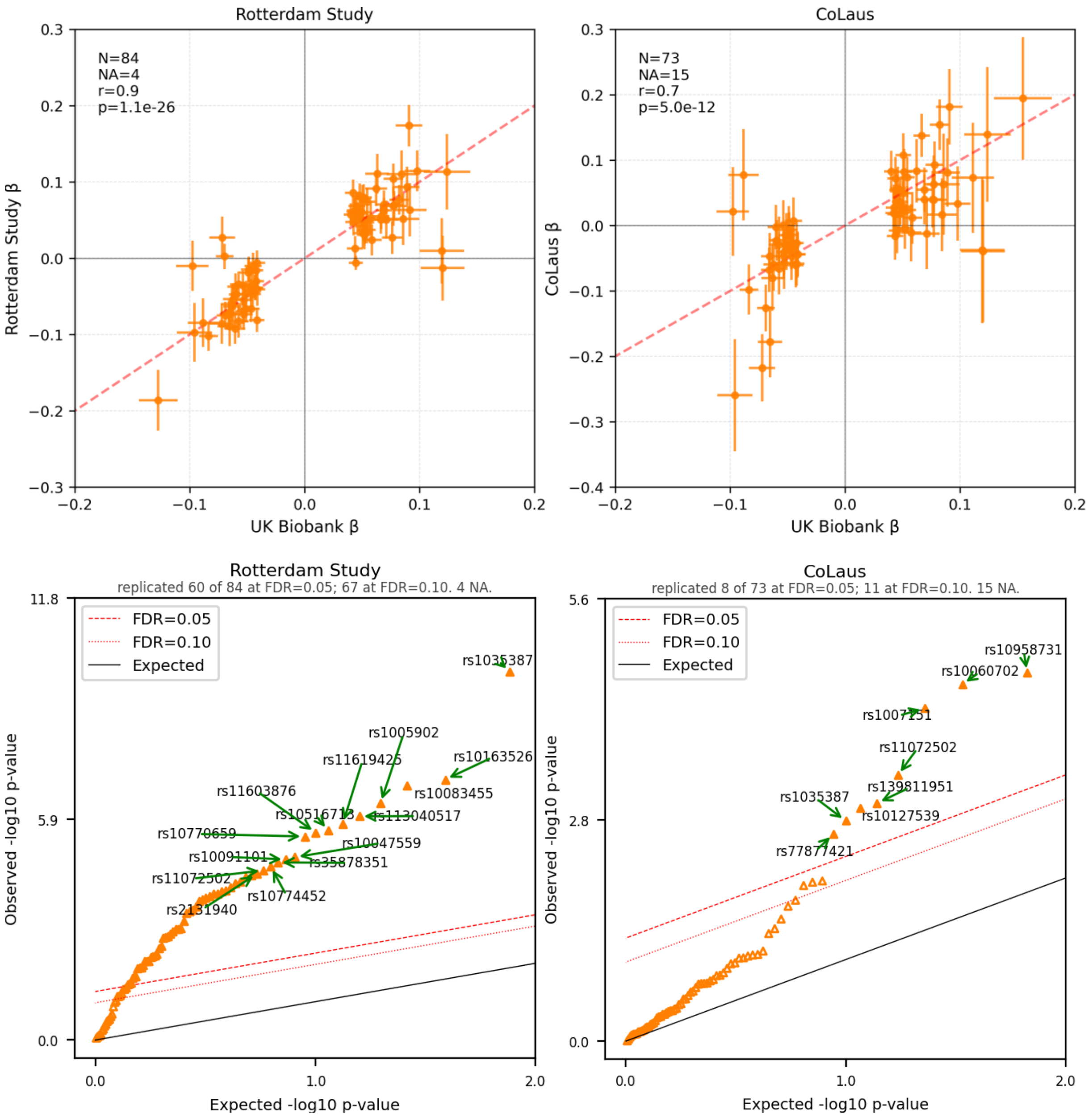

**Supplementary Figure 8 | Joint candidate SNP replication across cohorts.** Top panels show effect size estimates (β) for genome-wide significant SNPs identified in the UK Biobank (x-axis) versus replication cohorts (y-axis; left: Rotterdam Study, right: OphtalmoLaus / CoLaus). Bottom panels show corresponding quantile–quantile (QQ) plots of SNP-level p-values in the replication cohorts. Black diagonal indicates the null expectation. The dotted red line indicates the FDR=0.1 threshold; the dashed red line indicates the FDR=0.05 threshold. The top 15 SNPs significant at a false discovery rate (FDR) of 5% are highlighted. Candidates were SNPs that achieved genome-wide significance in the UK Biobank.

#### 5.4 Joint candidate genes replication

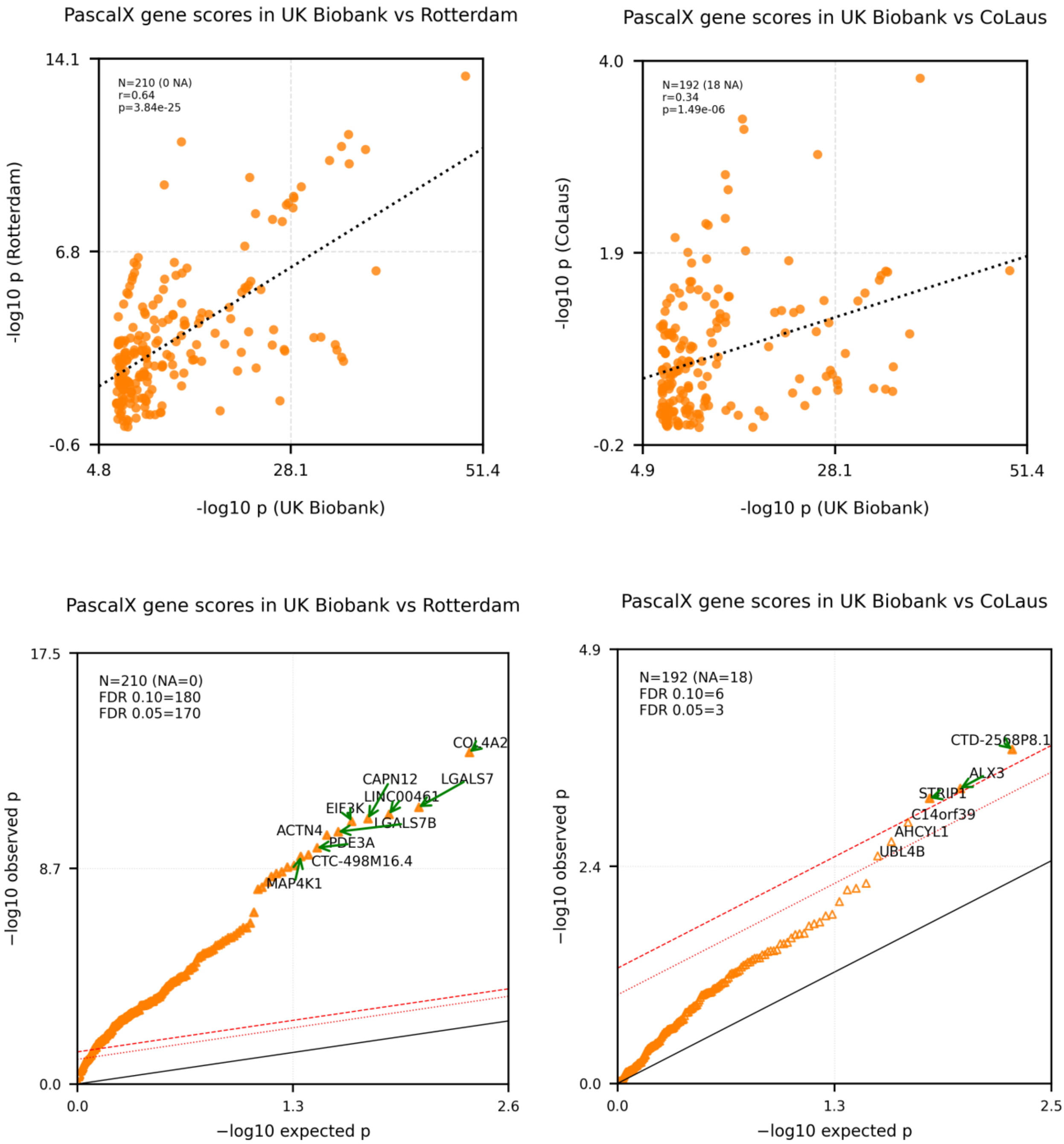

**Supplementary Figure 9 | Joint candidate gene replication across cohorts.** Gene-level association results obtained using *PascalX* in the UK Biobank (x-axis) versus replication cohorts (y-axis; left: Rotterdam Study, right: Ophtalmolaus / CoLaus). Top panels show scatterplots of −log10(p-values) per gene. Bottom panels show corresponding quantile–quantile (QQ) plots. Black diagonal indicates the null expectation. The dotted red line indicates the FDR=0.1 threshold; the dashed red line indicates the FDR=0.05 threshold. Top 10 genes significant at a false discovery rate (FDR) of 5% are annotated. Candidates were genes that achieved genome-wide significance in the UK Biobank.

#### 5.5 Trait-wise candidate replication in the Rotterdam Study

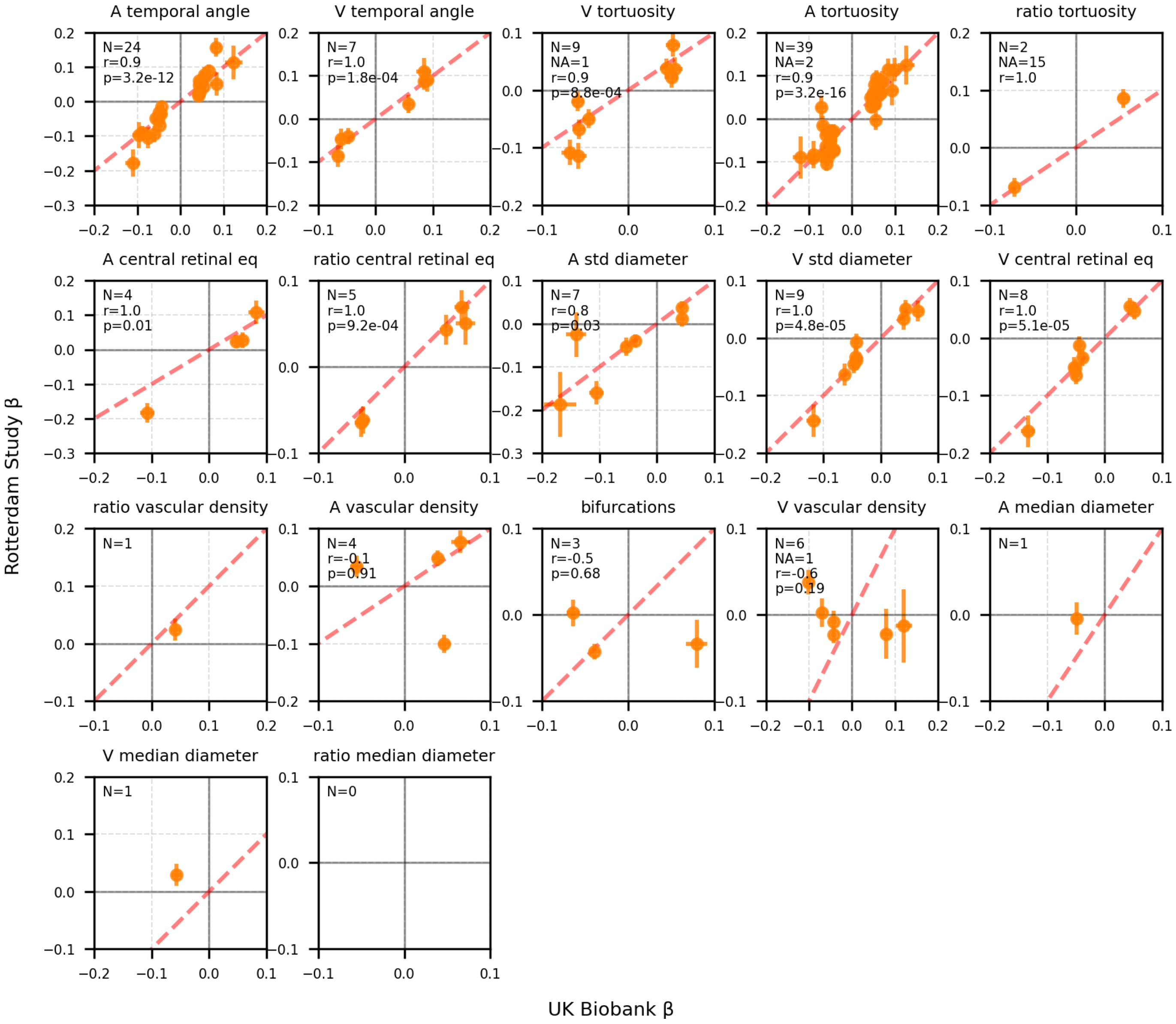

**Supplementary Figure 10 | Per-TIF SNP-level candidate replication in the Rotterdam Study.** Scatterplots comparing SNP-level association strengths (−log10 p-values) between the UK Biobank (x-axis) and the Rotterdam Study (y-axis) for candidate variants identified in the UK Biobank, shown separately for each dTIF. Each panel corresponds to one dTIF. Only candidate SNPs carried forward from the UK Biobank discovery analysis are shown. Pearson correlation coefficients and corresponding p-values are reported where applicable.

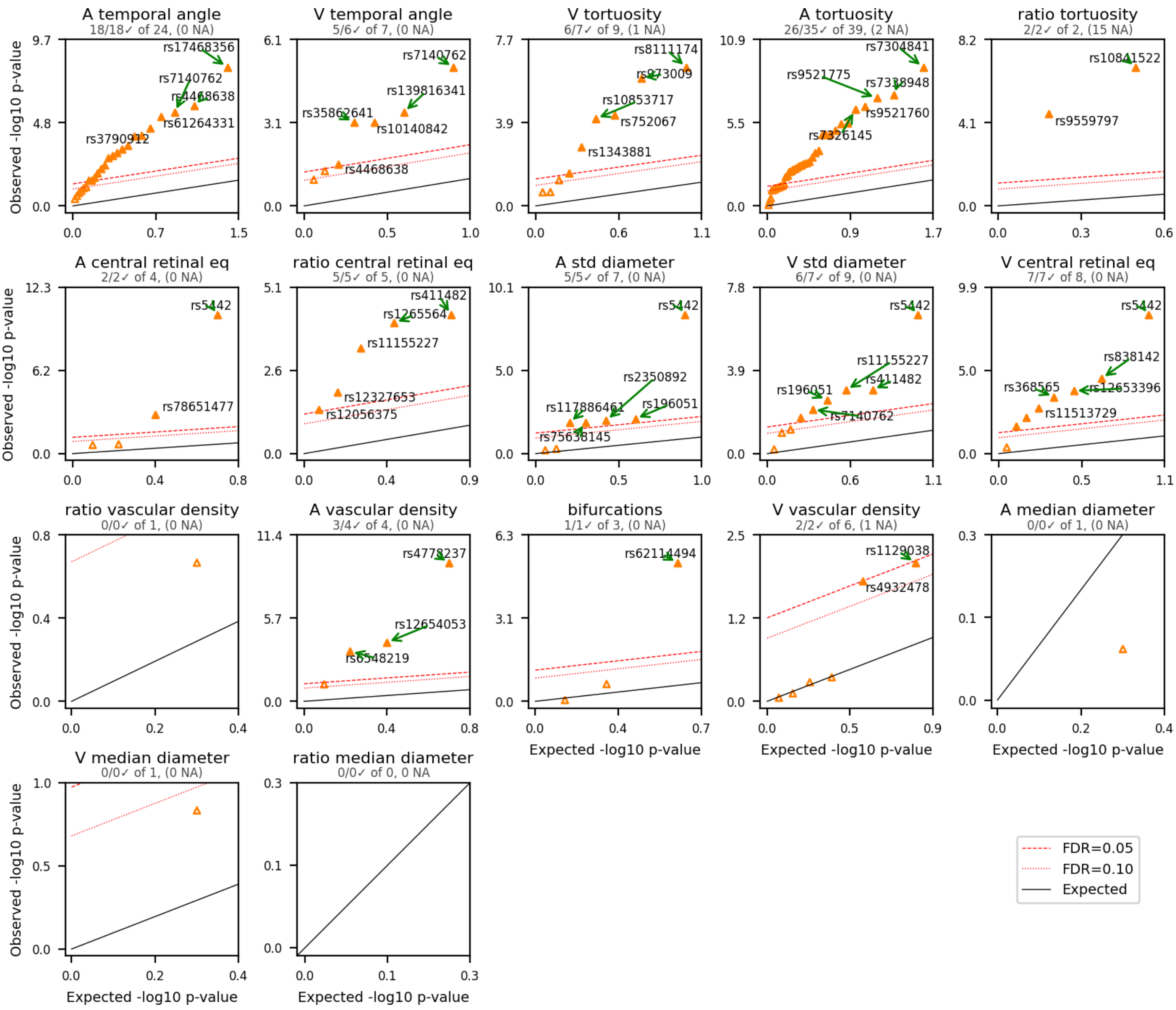

**Supplementary Figure 11 | Per-TIF SNP-level candidate association results in the Rotterdam Study.** Quantile–quantile (QQ) plots of SNP-level association p-values for each dTIF in the Rotterdam Study. Observed −log10(p-values) are plotted against the expected null distribution. Black diagonal indicates the null expectation. The dotted red line indicates the FDR=0.1 threshold; the dashed red line indicates the FDR=0.05 threshold. Top 5 SNPs significant at a Benjamini–Hochberg false discovery rate (FDR) of 5% are highlighted and annotated where applicable.

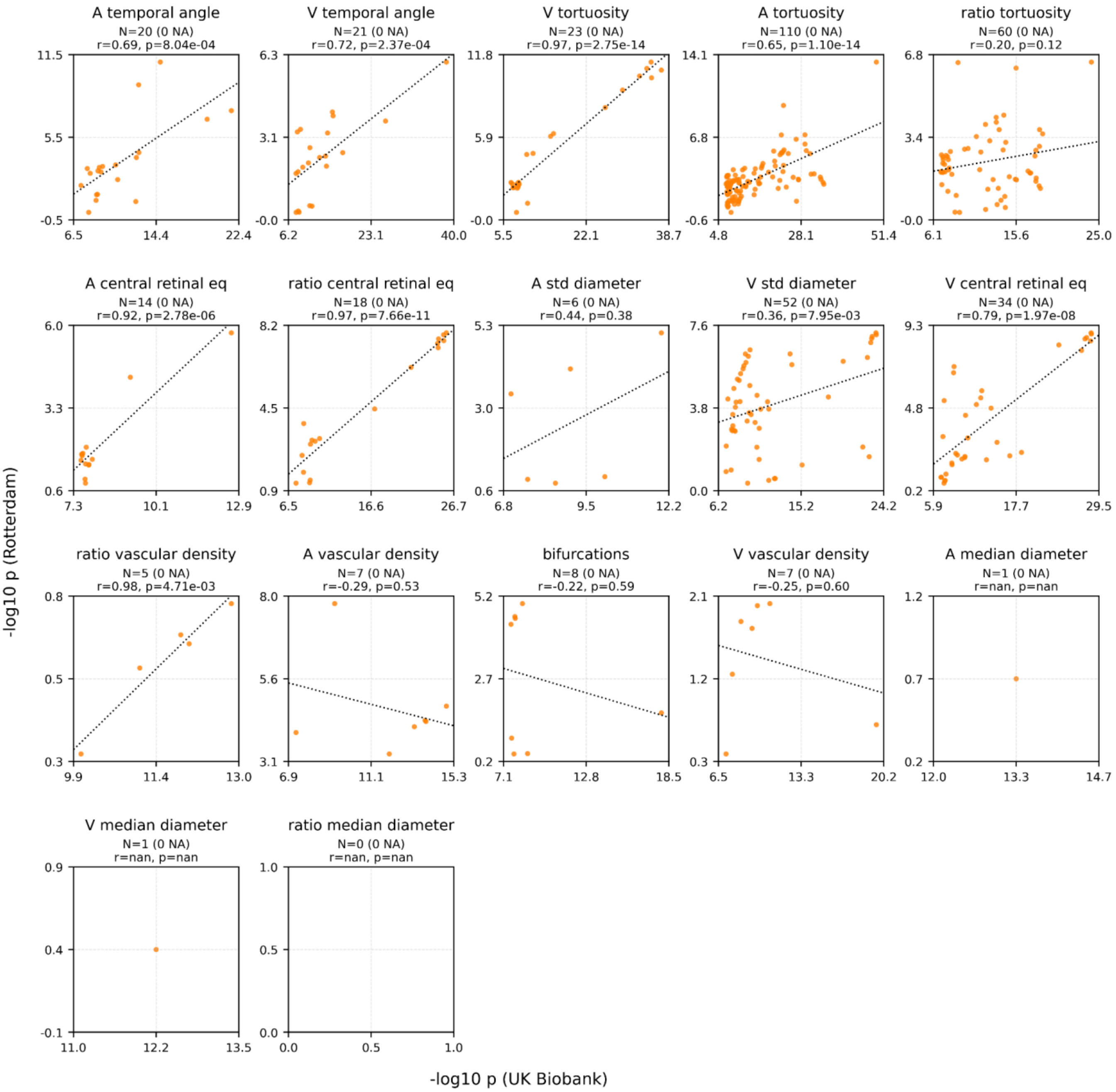

**Supplementary Figure 12 | Per-TIF candidate gene p-value correlation in the Rotterdam Study.** Scatterplots comparing gene-level association strengths (−log10 p-values) between the UK Biobank (x-axis) and the Rotterdam Study (y-axis) for each dTIF. Gene-level associations were computed using *PascalX*. Only candidate genes (genome-wide significant in the UK Biobank discovery analysis) are shown. Each panel corresponds to one dTIF; each point represents one gene. Pearson correlation coefficients and corresponding p-values are shown where applicable.

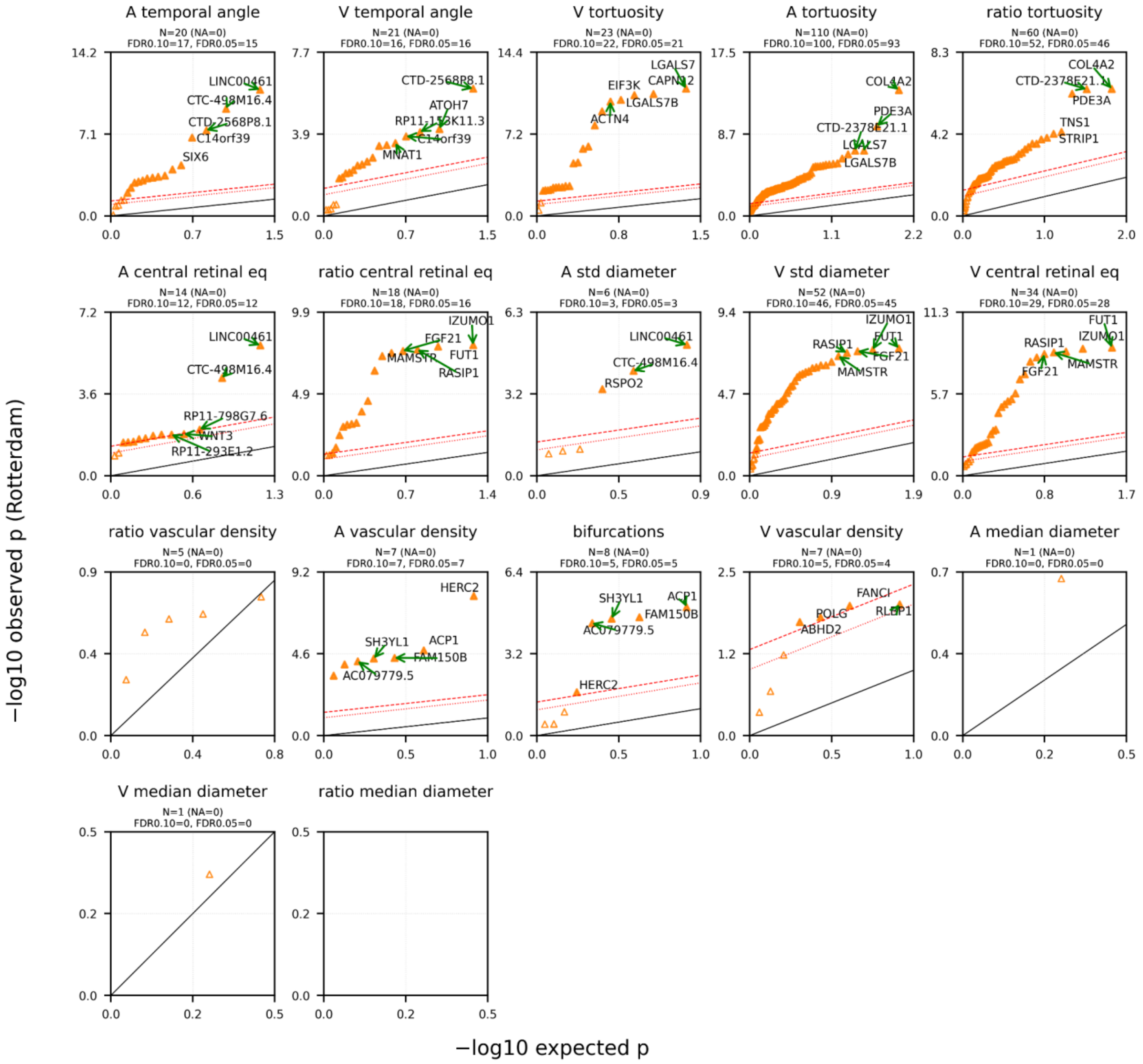

**Supplementary Figure 13 | Per-TIF candidate gene replication in the Rotterdam Study.** QQ plots of gene-level association p-values for each dTIF in the Rotterdam Study. Gene-level associations were computed using *PascalX*. Only candidate genes (genome-wide significant in the UK Biobank discovery analysis) were tested. Observed −log10(p-values) are plotted against the expected null distribution. The black diagonal indicates the null expectation. The dotted red line indicates the FDR=0.1 threshold; the dashed red line indicates the FDR=0.05 threshold. Top 5 genes significant at a Benjamini–Hochberg false discovery rate (FDR) of 5% are highlighted and annotated where applicable.

#### 5.6 Trait-wise candidate replication in OphtalmoLaus

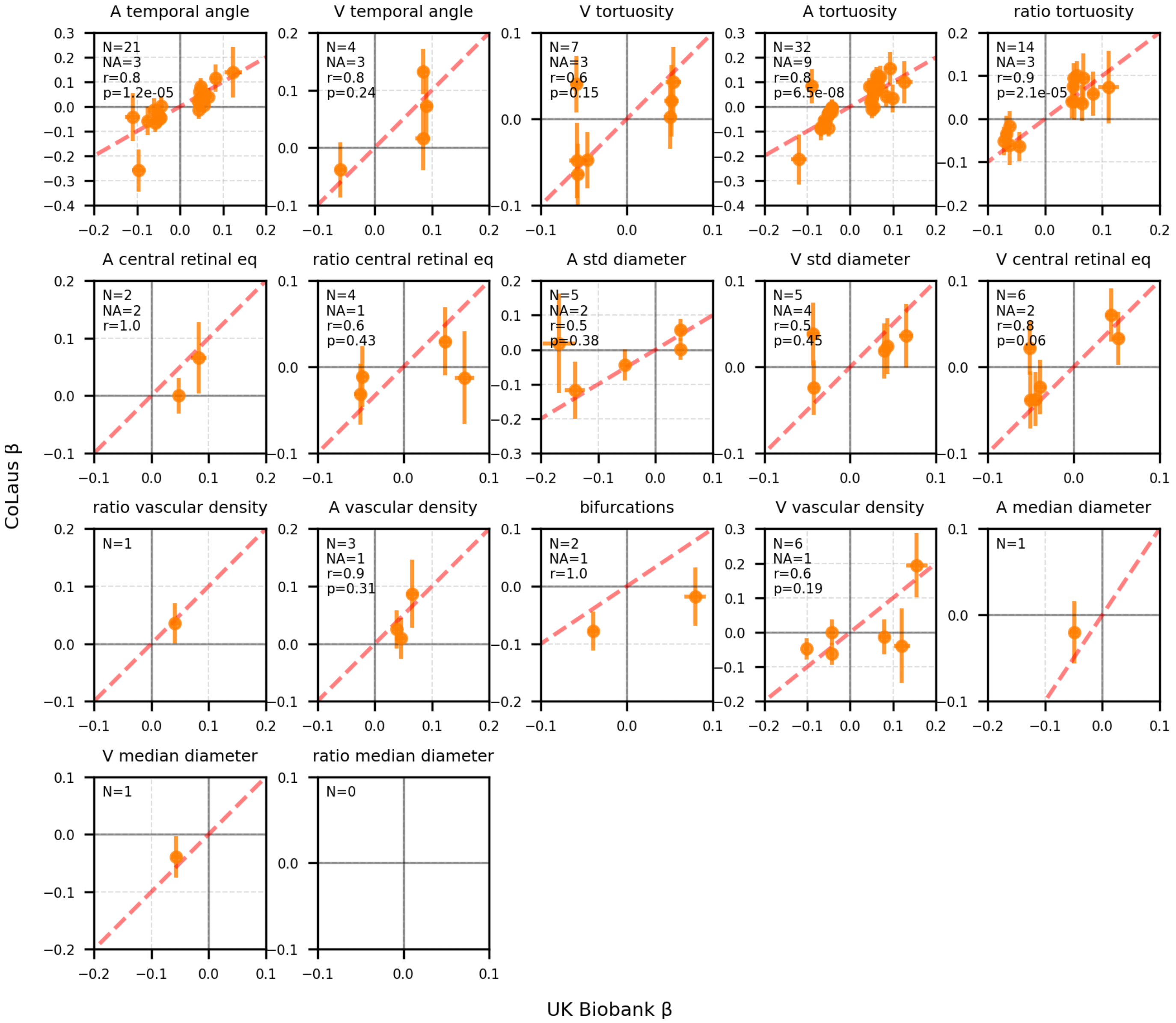

**Supplementary Figure 14 | Per-TIF SNP-level candidate replication in the OphtalmoLaus.** Scatterplots comparing SNP-level association strengths (−log10 p-values) between the UK Biobank (x-axis) and OphtalmoLaus (CoLaus; y-axis) for candidate variants identified in the UK Biobank, shown separately for each dTIF. Each panel corresponds to one dTIF. Only candidate SNPs carried forward from the UK Biobank discovery analysis are shown. Pearson correlation coefficients and corresponding p-values are reported where applicable.

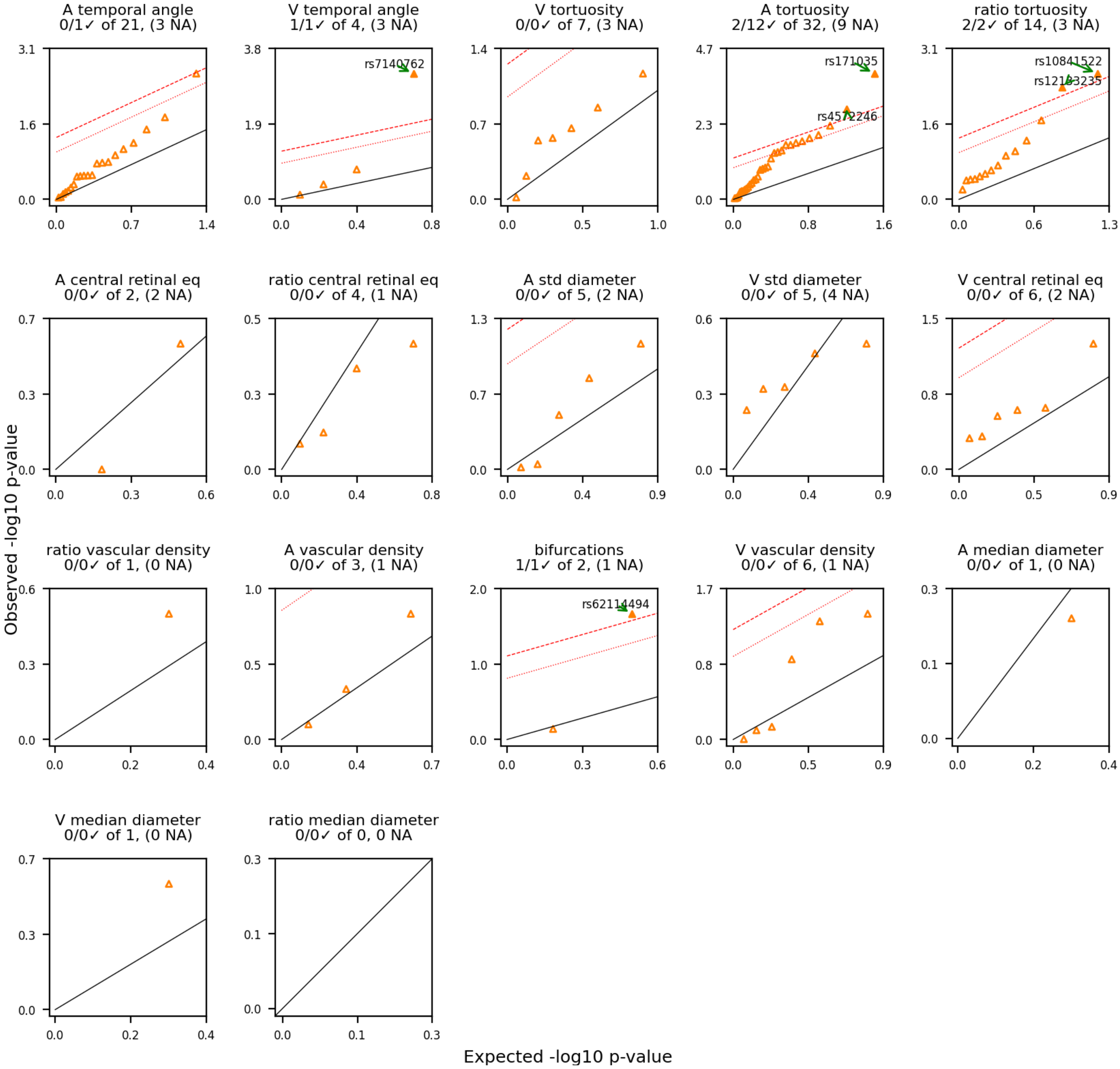

**Supplementary Figure 15 | Per-TIF SNP-level candidate association results in the OphtalmoLaus.** Quantile–quantile (QQ) plots of SNP-level association p-values for each dTIF in the OphtalmoLaus (CoLaus). Observed −log10(p-values) are plotted against the expected null distribution. Black diagonal indicates the null expectation. The dotted red line indicates the FDR=0.1 threshold; the dashed red line indicates the FDR=0.05 threshold. Top 5 SNPs significant at a Benjamini–Hochberg false discovery rate (FDR) of 5% are highlighted and annotated where applicable.

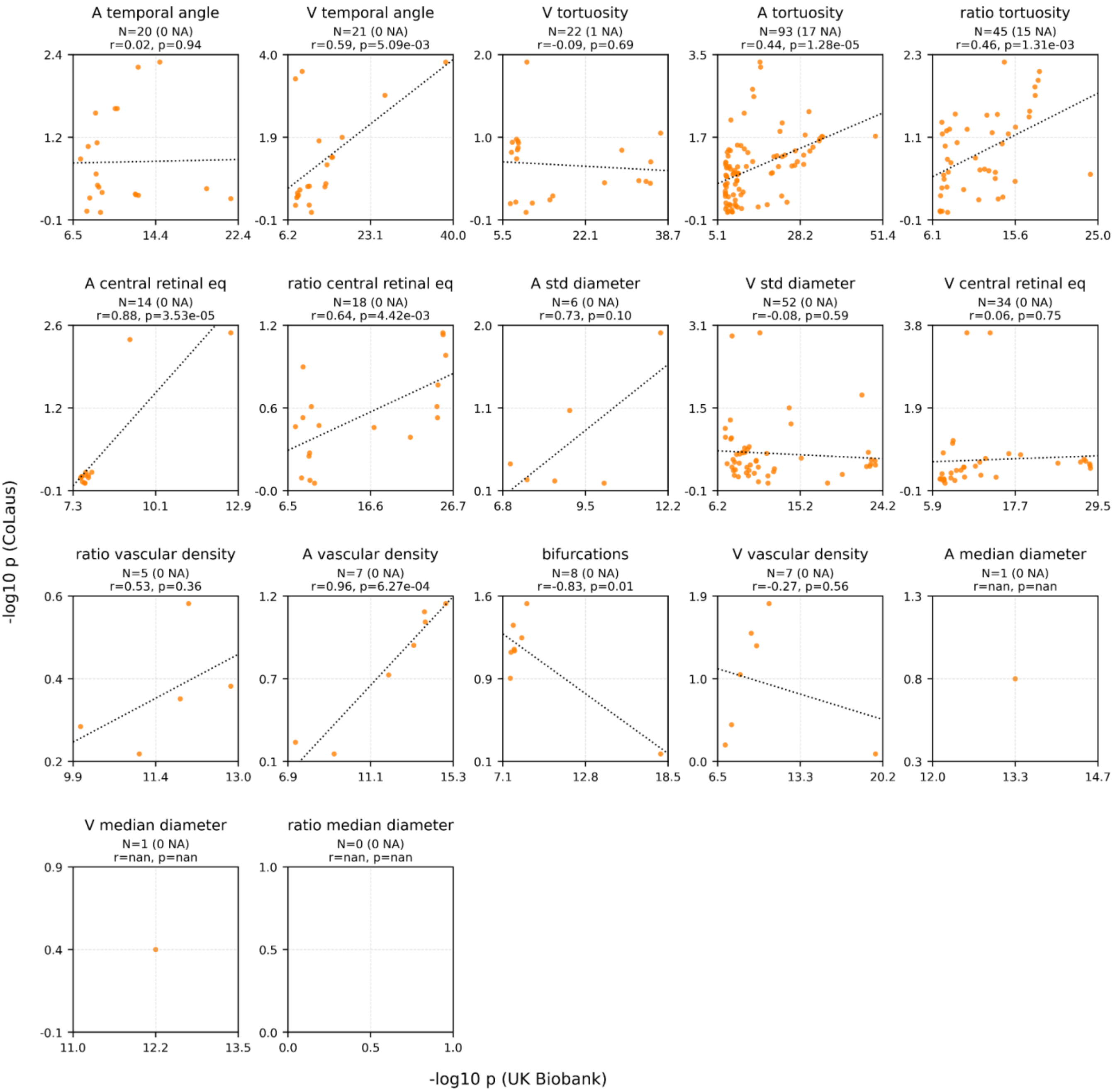

**Supplementary Figure 16 | Per-TIF candidate gene p-value correlation in the OphtalmoLaus.** Scatterplots comparing gene-level association strengths (−log10 p-values) between the UK Biobank (x-axis) and OphtalmoLaus (CoLaus; y-axis) for each dTIF. Gene-level associations were computed using *PascalX*. Only candidate genes (genome-wide significant in the UK Biobank discovery analysis) are shown. Each panel corresponds to one dTIF; each point represents one gene. Pearson correlation coefficients and corresponding p-values are shown where applicable.

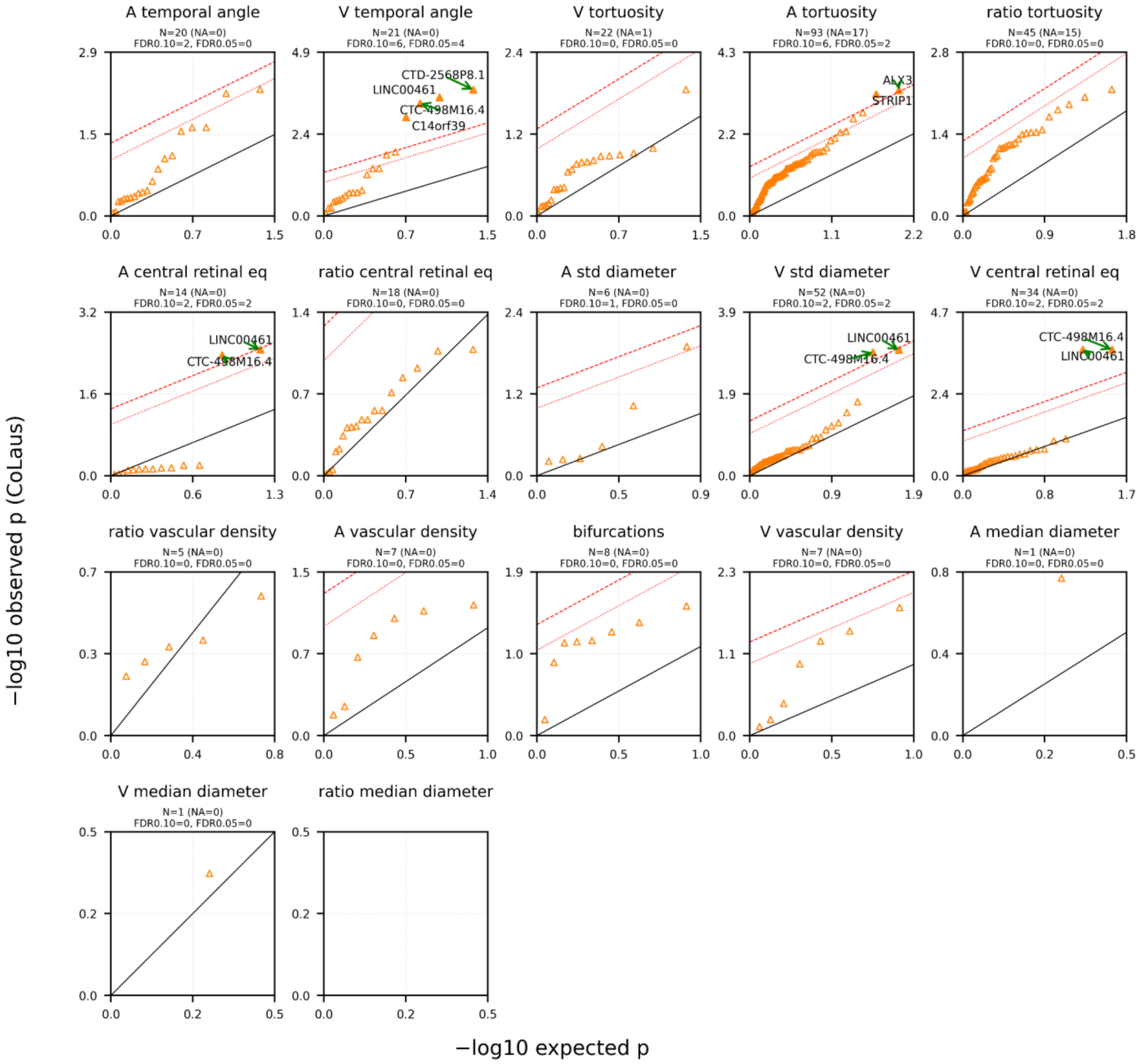

**Supplementary Figure 17 | Per-TIF candidate gene replication in the OphtalmoLaus.** QQ plots of gene-level association p-values for each dTIF in OphtalmoLaus (CoLaus). Gene-level associations were computed using *PascalX*. Only candidate genes (genome-wide significant in the UK Biobank discovery analysis) were tested. Observed −log10(p-values) are plotted against the expected null distribution. The black diagonal indicates the null expectation. The dotted red line indicates the FDR=0.1 threshold; the dashed red line indicates the FDR=0.05 threshold. Top 5 genes significant at a Benjamini–Hochberg false discovery rate (FDR) of 5% are highlighted and annotated where applicable.

#### 5.7 Distribution of mTIF and dTIF heritabilities in the Rotterdam Study

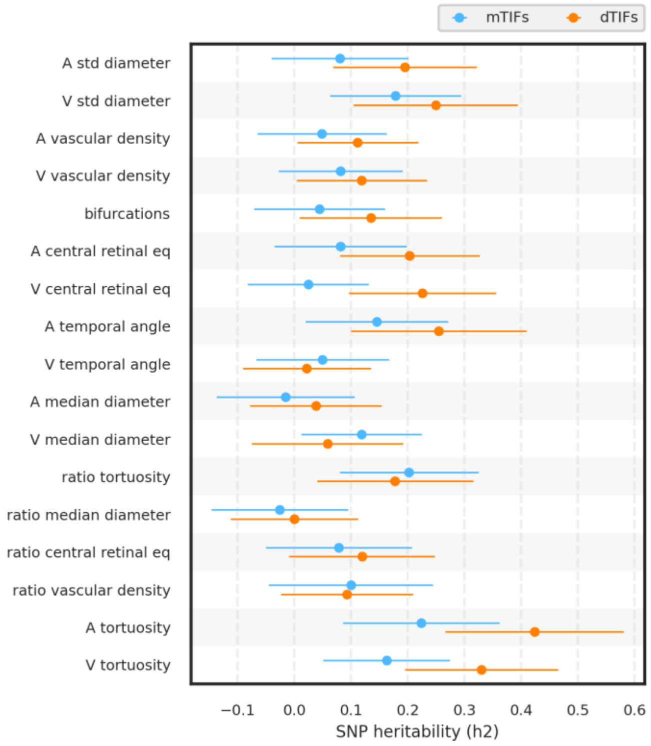

**Supplementary Figure 18 | Forest plot of SNP heritabilities (h2) in the Rotterdam Study for mTIFs and dTIFs.** Points show *LDSR* SNP-wise heritabilities ($h^2$) with 95% CI (±1.96 SE). Inverse-variance–weighted paired comparison (dTIF − mTIF) across matched traits (n=17) indicated a significant increase in heritability in dTIFs compared to mTIFs: mean difference=0.066 (SE 0.0215), z=3.08, p=0.0021, 95% CI [0.024, 0.109], Cohen's d=0.86. CI: confidence interval, SE: standard error

#### 5.8 Replication of disease associations in Rotterdam Study

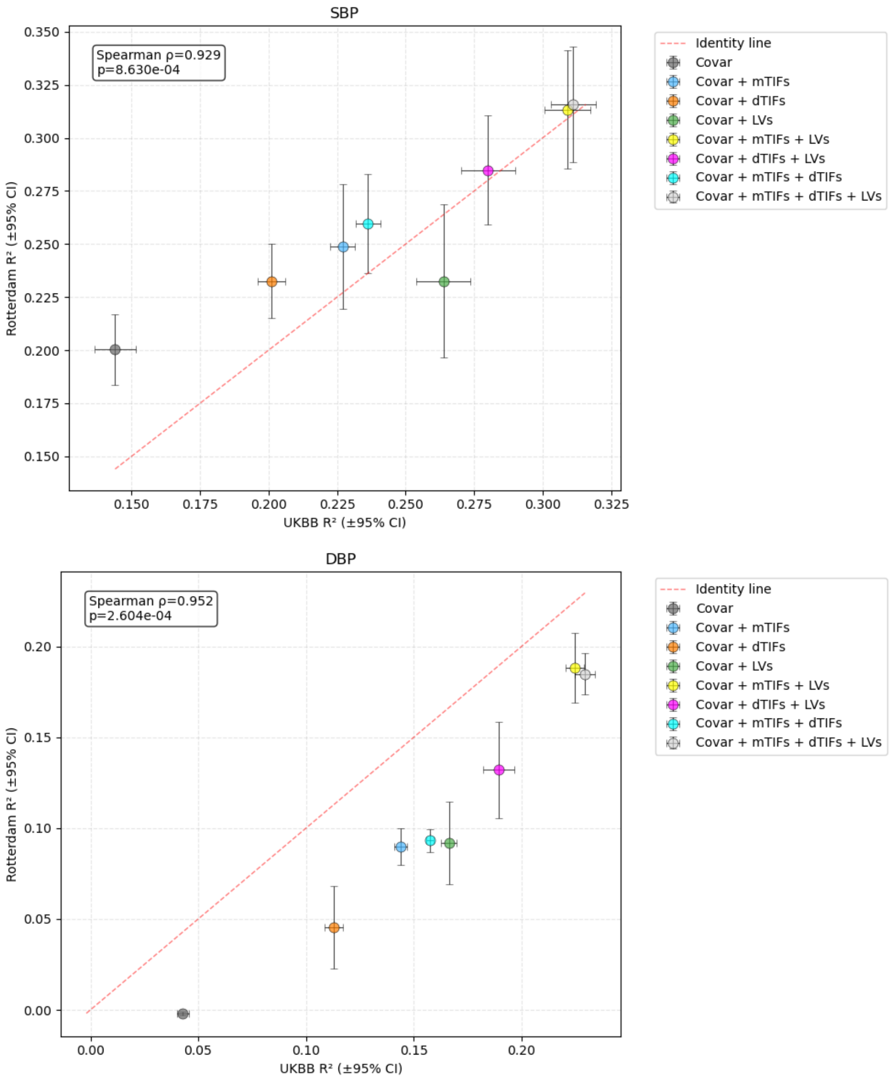

**Supplementary Figure 19 | Correlations of predictive power ($R^2$) between UK Biobank (UKBB) and Rotterdam Study.** Regularized linear models to predict systolic blood pressure (SBP) and diastolic blood pressure (DBP). Feature sets include: (i) a baseline model including only covariates (see methods for list), (ii) all 17 mTIFs, (iii) all 17 dTIFs, (iv) 1024 RETFound LVs, (v) all mTIF and 1024 RETFound LVs, (vi) all dTIFs and 1024 RETFound LVs, (vii) all mTIFs and dTIFs, (viii) all features combined. Covar = Covariates. Error bars reflect 95% confidence intervals.

#### 6. Image quality distribution

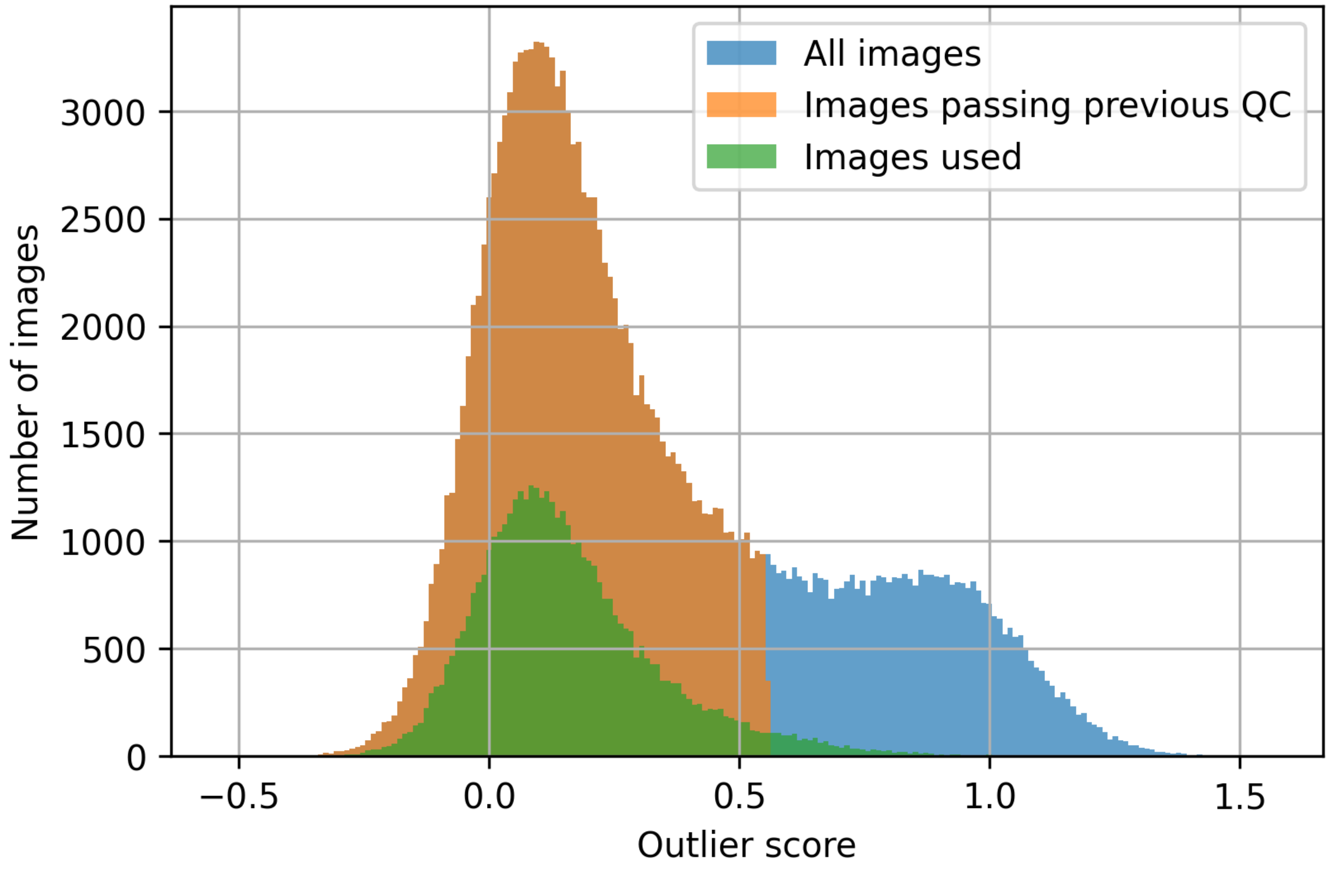

**Supplementary Figure 20 | Image quality distribution.** Image quality distribution in the UK Biobank according to a previously published quality assessment method [1]. The "outlier score" is higher for images of lower quality.

#### 7. Distributions of covariates

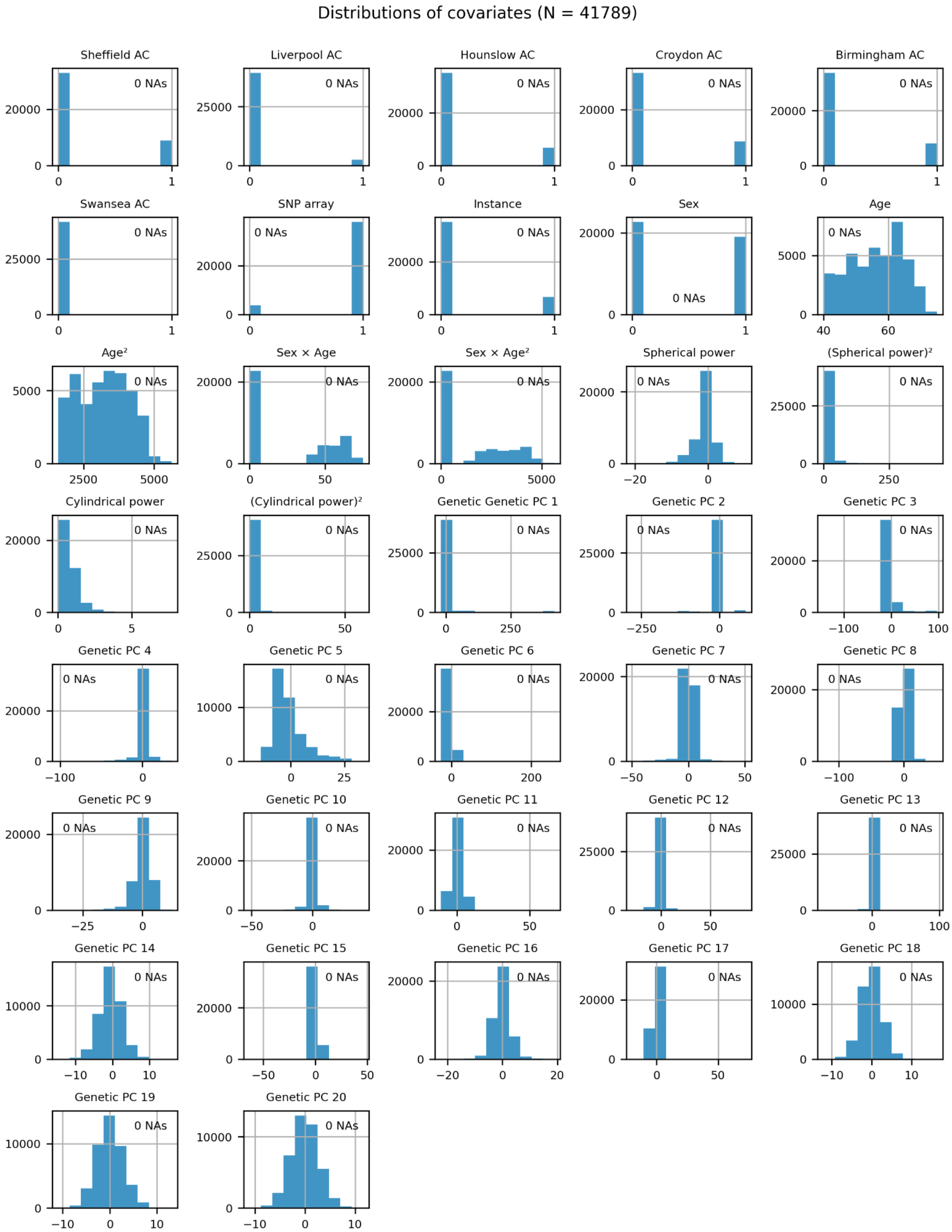

**Supplementary Figure 21 | Distribution of covariates.** Raw distributions of covariates that were used to correct TIFs and LVs in our genetic analyses and disease associations. No covariates had any missingness.

### 8. Distributions of diseases and risk factors

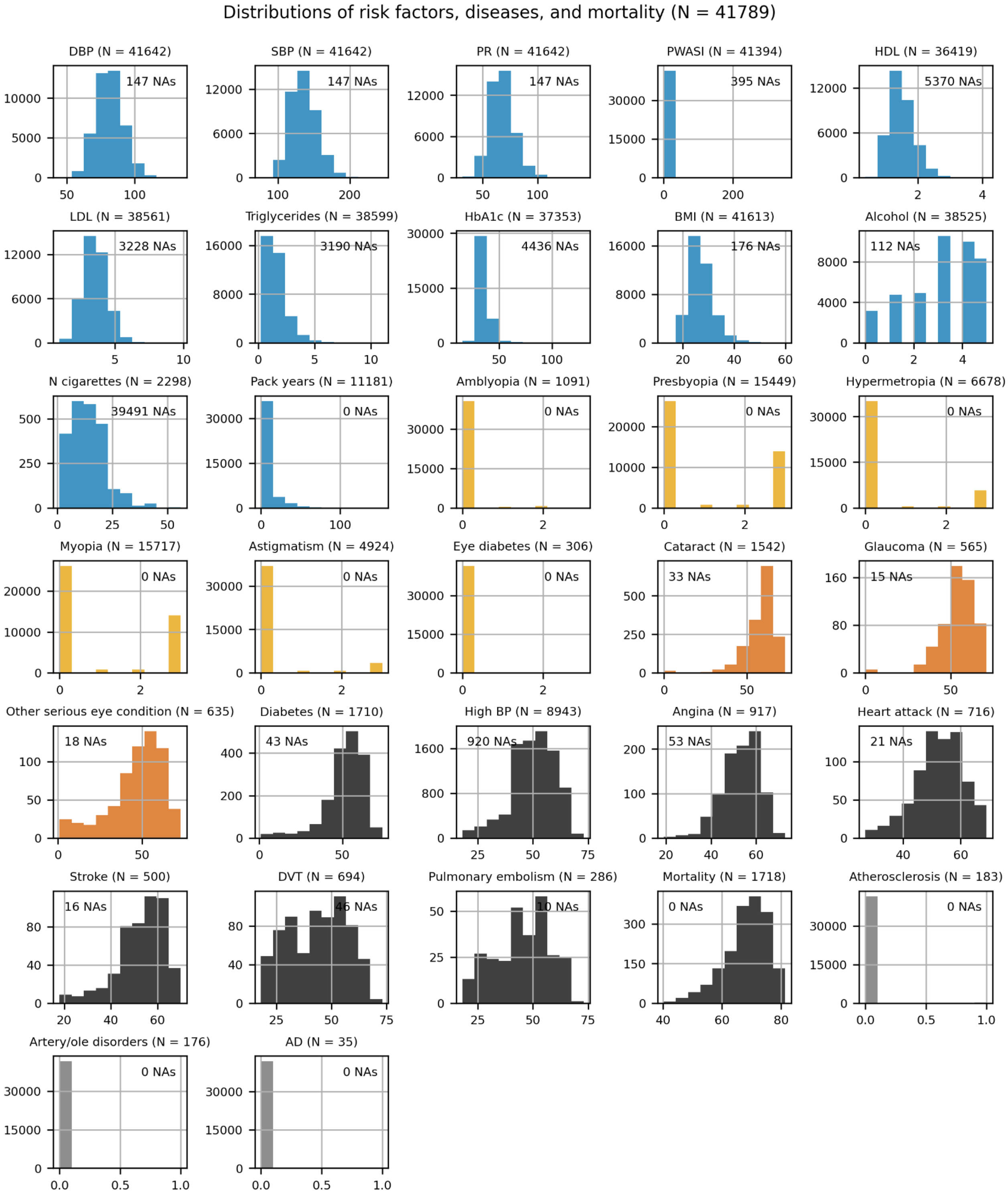

**Supplementary Figure 22 | Distribution of diseases.** Blue: risk factors, orange: ocular diseases, dark orange: ocular diseases with age-at-onset data, dark gray: systemic diseases with age-at-onset data and mortality, light gray: binary systemic diseases.
